# Retrospective evaluation of school-related measures on pre-vaccination transmission dynamics of SARS-CoV-2

**DOI:** 10.1101/2025.04.14.25325720

**Authors:** Benedetta Canfora, Rey Audie Escosio, Christiaan H. van Dorp, Otilia Boldea, Marc Bonten, Ana Nunes, Ganna Rozhnova

## Abstract

Schools are important settings for respiratory virus transmission and a major focus of pandemic control measures in the absence of effective pharmaceutical interventions. During the COVID-19 pandemic, decisions regarding school closures and openings were highly debated due to uncertainties about their population-level impact. Using an age-stratified transmission model formally fitted to hospitalization and seroprevalence data in a Bayesian framework, we retrospectively assessed the effect of school closures and openings on SARS-CoV-2 transmission in Portugal before mass vaccination. We compared the observed epidemic trajectory with two counterfactual scenarios: one in which schools remained open during the first lockdown of spring 2020 and another in which they closed for in-person education following the summer holidays in autumn 2020. We found that keeping schools open during the first lockdown could have led to a worse situation, with hospitalizations rising far beyond observed levels in spring 2020. However, closing schools after the summer holidays alone would not have been sufficient to prevent the autumn wave of hospitalizations in 2020, though it could have mitigated its impact. These findings underscore that the epidemiological impact of school closures is highly context-dependent, influenced by mitigation measures within schools and the intensity of transmission outside them. While closures can support control efforts, especially early into a pandemic, their benefits must be weighed against substantial societal costs. Our results suggest that integrated strategies combining school-based and broader community interventions are essential to effectively reduce transmission during future pandemics.

## Introduction

The COVID-19 pandemic prompted governments worldwide to implement a range of public health measures to control the spread of SARS-CoV-2, with schools becoming a primary target for interventions. Given the absence of mass vaccination in the first pandemic year, school-based interventions were particularly widespread, making it crucial to assess their role in reducing transmission. In Portugal, school-related measures were guided by recommendations from the Directorate General of Health [1] and included key actions aimed at reducing transmission-relevant contacts, such as reduced classroom capacity, increased outdoor activities, staggered schedules, and stringent cleaning protocols. Portuguese schools closed in March 2020, shortly after the detection of the initial SARS-CoV-2 cases, and all educational activities were suspended until mid-April 2020 [2, 3]. Following a temporary relaxation of restrictions, schools partially reopened in late May 2020 [1]. By September 2020, all students had returned to in-person learning [4] and schools remained open until the end of the year [5, 6].

The impact of school-based measures on controlling the pandemic was extensively studied across different countries [7–13], but findings varied depending on the context and the concurrent implementation of other public health measures in the community. While school closures were among the most widely used interventions, studies suggested that this measure alone was insufficient to control community transmission [9, 10]. For instance, studies for the Netherlands and the United Kingdom demonstrated that the effectiveness of school closures depended heavily on the context and the availability of non-school-based interventions [8, 11]. Moreover, while reducing school-based contacts likely curtailed in-school transmission, this effect may have been offset by increased household interactions, potentially diminishing their effectiveness [14]. Consequently, decisions regarding school closures required a careful balance between epidemiological benefits and their substantial educational, social, and economic costs [12].

Despite extensive research on school-based interventions in other countries, their impact within the Portuguese context remains underexplored. Most SARS-CoV-2 transmission models for Portugal primarily focused on epidemiological profiling [15–17], parameter estimation [2, 18–20], short-term predictions [21], and the evaluation of interventions such as vaccination [5,22,23], lockdowns, quarantines [23–25], and combinations thereof [26–28]. Some studies indirectly considered school-based interventions within broader lockdown scenarios [22, 25], but few studies explicitly examined their effects through statistical [4] or deterministic compartmental models [6, 29]. These studies focused on predictive modeling of school-based interventions on the pandemic waves in 2021, whereas a retrospective evaluation of their impact during the pre-vaccination period, a time of stringent government interventions and heightened public health response, has not been performed. Furthermore, modeling school-based interventions requires accurate estimation of age-specific, time-varying parameters (e.g., susceptibility and contact patterns), yet these factors have been largely overlooked in previous studies. Filling these gaps is essential for retrospectively assessing the role of schools in transmission during the early pandemic waves before mass vaccination when non-pharmaceutical interventions played a crucial role.

This study aimed to retrospectively evaluate the impact of schools on SARS-CoV-2 transmission during the pre-vaccination period in Portugal. Using an age-stratified SARS-CoV-2 transmission model fitted to data, we assessed how different school closure and reopening strategies influenced COVID-19 hospital admissions and SARS-CoV-2 infections. Additionally, we estimated contact rates, effective reproduction numbers, and the relative contributions of different age groups to transmission, providing insights into key drivers of viral spread before the onset of mass vaccination in Portugal.

## Data and methods

### Data

To parameterize and train our transmission model, we used publicly available data on SARS-CoV-2 seroprevalence, COVID-19 hospital admissions, demographics, and contact patterns. The national age-specific seroprevalence data, including sample size, number of positive samples, and 95% confidence intervals stratified by age group, were obtained from the First National Serological Survey (ISNCOVID-19) conducted in Portugal between 21 May 2020 and 8 July 2020 [30]. The national age-specific COVID-19 hospital admission data from 2 March 2020 to 15 January 2021 were provided by the Central Administration of the Health System and the Shared Services of the Ministry of Health. The national age-specific demographic data for Portugal in 2019 were obtained from the Contemporary Portugal Database [31]. The pre-pandemic contact matrices for school-based and total transmission-relevant contacts in Portugal were taken from Mistry et al [32]. Due to the lack of contact data for Portugal during the first lockdown, such as that provided by Mistry et al [32] for the pre-pandemic period, we followed our previous approach [23] and inferred the Portuguese contact matrix for this period using data from the Netherlands [33], where a comparable set of interventions was implemented.

### Transmission model

This work builds upon an age-stratified SARS-CoV-2 transmission model in [23] that was formulated as a system of ordinary differential equations for the number of individuals in age group *k*, *k* = 1*, … , n*, who are susceptible (*S_k_*), exposed (*E_k_*), infectious (*I_k_*), recovered (*R_k_*), and hospitalized (*H_k_*) at time *t*. The model equations are given by

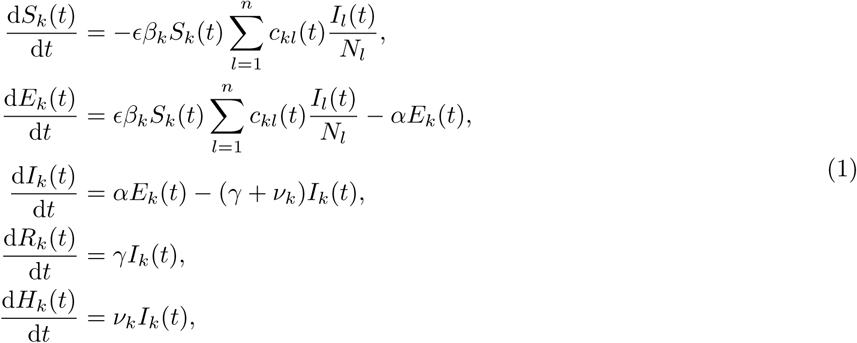

where *N_k_*is the number of individuals in age group *k*, *N_k_*= *S_k_*(*t*) + *E_k_*(*t*) + *I_k_*(*t*) + *H_k_*(*t*) + *R_k_*(*t*).

This deterministic compartmental model followed susceptible-exposed-infectious-recovered-hospitalized dynamics. Susceptible individuals in age group *k* (*S_k_*) become latently infected (*E_k_*) through contacts with infectious individuals in age group *l* (*I_l_*) occurring at a rate *c_kl_*(*t*). The probability of transmission per contact is *ɛ*, and the relative susceptibility of individuals in age group *k* is *β_k_*. Exposed individuals (*E_k_*) become infectious (*I_k_*) at a rate *α*. Infectious individuals (*I_k_*) either become hospitalized at an age-specific rate *ν_k_* or recover without hospitalization at a rate *γ*. To account for age-related differences in parameters, the population was stratified into *n* = 10 age groups, indexed by *k* (*k* = 1*, … , n*): [0, 5), [5, 10), [10, 20), [20, 30), [30, 40), [40, 50), [50, 60), [60, 70), [70, 80), and 80+ years. The simulations began at *t* = 0, corresponding to February 26, 2020, when a fraction *θ* of the population was already infected. This fraction was assumed to be evenly distributed between the exposed (*E_k_*) and infectious (*I_k_*) compartments, while the remaining population was susceptible: 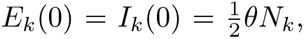 *S_k_*(0) = (1 − *θ*)*N_k_*, *R_k_*(0) = *H_k_*(0) = 0. A summary of all model parameters as defined in various sections of the Methods is given in Supplementary Table S1.

The model incorporated several simplifying assumptions. First, demographic dynamics and immunity waning were not considered due to the short timeframe of the analyses of less than one year. Second, hospitalized individuals were assumed to be non-infectious, as they constituted a small proportion of the total infectious population and were subject to various personal protective measures in hospitals. Third, since the model was fitted to hospitalization incidence, intra-hospital dynamics, and disease-related deaths were not explicitly modeled. Fourth, as hospitalization and seroprevalence data were only available at the national level, additional spatial stratification was not included. Lastly, since the period during which the B.1.1.7 variant was dominant falls outside the timeframe of analyses, this variant of concern was not explicitly modeled.

### Contact rates

#### Pandemic timeline

The schematic timeline of the pre-vaccination phase of the pandemic, spanning from 26 February 2020 to 15 January 2021, is shown in Figure 1. Following an initial stage without any interventions, various control measures were implemented, defining distinct regimes throughout 2020: (i) the first lockdown, during which schools were closed in late March; (ii) a relaxation of restrictions following the first lockdown, beginning in late May; (iii) further easing of measures, including the reopening of schools after the summer holidays in late August; (iv) the second lockdown in early November; and (v) a subsequent relaxation of measures in late December. These regimes were denoted by the index *j* = 0, 1, 2, 3, 4 in the analysis that follows.

**Figure 1.**
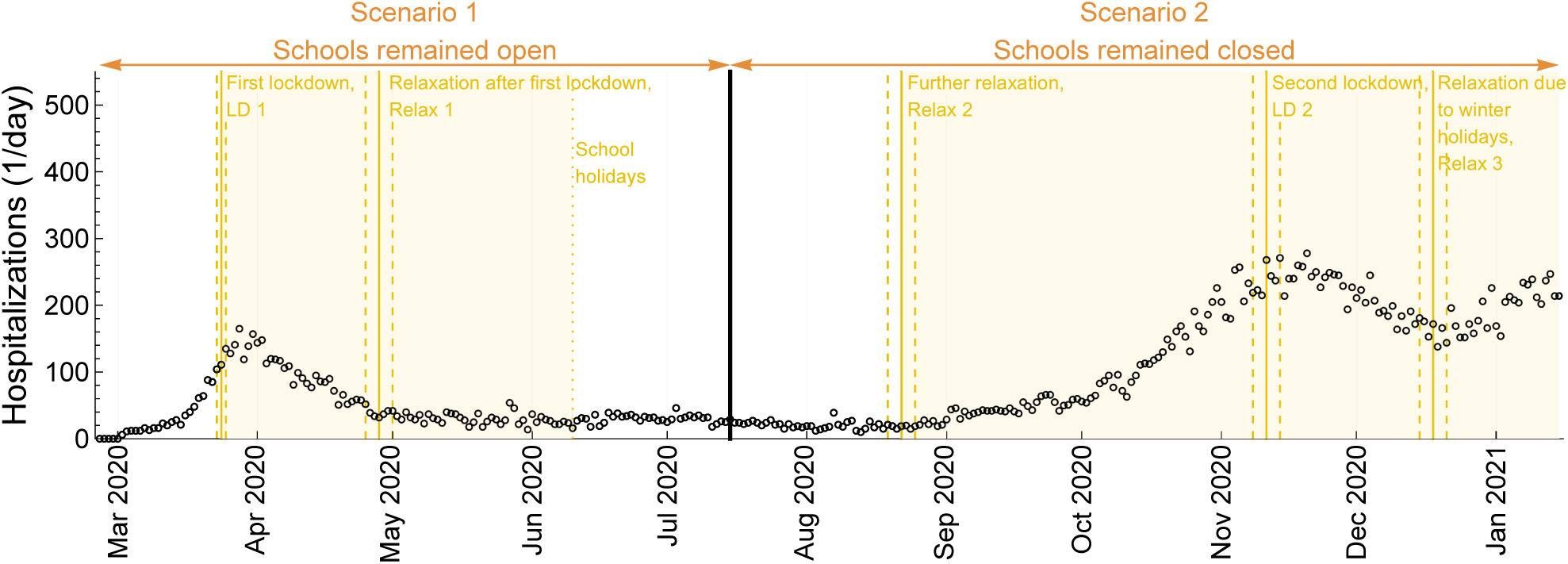
Schematic timeline of the pandemic before vaccination in Portugal. The circles represent daily hospital admission data. The timeframes for two counterfactual scenarios, indicated by the orange arrows at the top, are separated by the thick solid black line. The baseline refers to the real-world scenario. Scenario 1: schools remained open during LD 1 and Relax 1 until the summer school holidays starting on 10 June 2020 (Baseline: schools were closed during LD 1 and open during Relax 1). Scenario 2: schools remained closed after the 2020 summer holidays during Relax 2, LD 2, and Relax 3 (Baseline: schools were open during Relax 2, LD 2, and Relax 3). The yellow-shaded area represents the period when schools were open in Scenario 1 and closed in Scenario 2, respectively. The solid yellow lines, each described by their respective annotations, denote the median mid-point times of transition estimated from the model, while the enclosing dashed yellow lines indicate the 95% CrI. The dotted yellow line on 10 June 2020 marks the start of the school summer holidays. LD: lockdown. Relax: relaxation of measures.

#### Counterfactual scenarios vs baseline

We retrospectively evaluated two counterfactual scenarios (Figure 1): (i) Scenario 1, where schools remained open during the first lockdown (LD 1) and the first relaxation of measures (Relax 1) until summer school holidays in 2020 (Baseline: schools were closed during LD 1 and open during Relax 1), and (ii) Scenario 2, where schools remained closed after the 2020 summer holidays spanning the period of the second relaxation of measures (Relax 2), the second lockdown (LD 2), and the third relaxation of measures (Relax 3) (Baseline: schools were open during Relax 2, LD 2, and Relax 3). The yellow-shaded area in Figure 1 represents the period when schools were open in counterfactual Scenario 1 and closed in counterfactual Scenario 2, respectively. These scenarios are described in detail below. To distinguish these hypothetical scenarios from reality, we refer to the model fit to the observed data as the *baseline* or *real-world* scenario.

#### Time-varying contact patterns

The implementation of control measures leads to time-varying contact rates, *c_kl_*(*t*), which were estimated by fitting the model to observed data. The meaning of *c_kl_*(*t*) is the average number of daily transmission-relevant contacts (e.g., touching or a conversation) an individual in age group *k* makes with all individuals in age group *l*. Following the framework developed in [23], changes in contact rates were modeled as a series of smooth transitions between different regimes *j*, *j* = 0, 1, 2, 3, 4. For this, we used logistic functions, 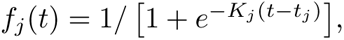 where *K_j_* is the speed of transition and *t_j_* is the mid-point of transition.

To define the contact rates *c_kl_*(*t*), we introduced the following notation. The pre-pandemic total and school-based contact rates were denoted by *b_kl_* and *s_kl_*, while *a_kl_* represents the contact rate during the first and most stringent lockdown. These contact matrices were obtained from the literature and served as data inputs for the model (Supplementary Figure S1). The parameter, 0 ≤ 1 − *ζ* ≤ 1, describes the reduction in transmission-relevant contacts during the first lockdown due to protective measures in the general population not explicitly modeled, such as mask-wearing, refraining from shaking hands, and physical distancing. Similarly, the parameter, 0 ≤ 1 − *ζ_s_* ≤ 1, describes the reduction in transmission-relevant school contacts due to protective measures among students and teachers. The parameters, 0 ≤ *u_j_* ≤ 1, where *j* = 1, 2, 3, 4, were introduced to model changes in contact behavior. These parameters can be interpreted as the average fraction of time individuals behaved as before the pandemic, as described by the contact matrix *b_kl_*, with (1 − *u_j_*) indicating the proportion of time they behaved as during the first and most stringent lockdown, as described by the contact matrix *ζa_kl_*. School closures for summer holidays, relevant in Scenario 1, were also modeled using a logistic function 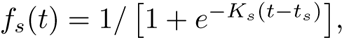 where *t_s_* corresponds to 10 June 2020 and, for simplicity, *K_s_* is chosen so that all schools close on the same day. A summary of the model parameters relating to contact patterns is given in Supplementary Table S1.

#### Scenario 1 versus baseline

For the period from 26 February to 15 July 2020, the baseline contact rate was estimated as follows

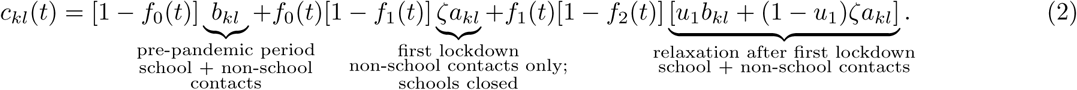

Eq. (2) implies that contact rates transitioned from *b_kl_* before the lockdown to *ζa_kl_* during the lockdown, governed by the function *f*_0_(*t*). For later time points, contact rates were modeled as a linear combination of *b_kl_* and *ζa_kl_*. Specifically, contact rates transitioned from *ζa_kl_* in the lockdown to [*u*_1_*b_kl_* + (1 − *u*_1_)*ζa_kl_*] during the relaxation period, governed by the function *f*_1_(*t*). Subsequent contact modeling followed a similar approach.

In the same period, the contact rate for Scenario 1, where schools remained open instead of closing during the first lockdown, was modeled as

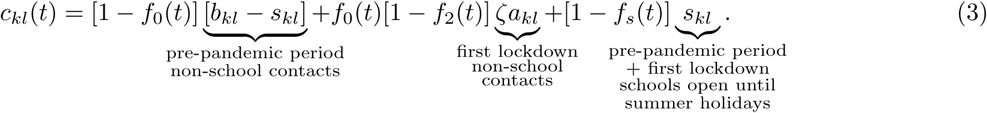

In Eq. (3), we assumed that (i) the total contact rate in the pre-pandemic period, *b_kl_*, was the sum of school contacts, *s_kl_*, and non-school contacts, (*b_kl_* − *s_kl_*) [32]; (ii) schools stayed open during the first lockdown and the first relaxation of measures and closed for summer holidays on 10 June 2020; (iii) since keeping schools open resulted in a hospitalization wave larger than the first pandemic wave, the relaxation of measures affecting non-school contacts was not implemented until hospitalizations declined to the level at which the first lockdown was relaxed. Consequently, the last term in Eq. (2) was omitted from Eq. (3). It is important to emphasize that assumption (iii), which prevented the relaxation of measures in late May amid a surge of hospitalizations, was introduced to maintain the plausibility of our retrospective Scenario 1. Results without this assumption are presented in the sensitivity analyses section below.

#### Scenario 2 versus baseline

For the period from 15 July 2020 to 16 January 2021, the baseline contact rate was estimated as follows

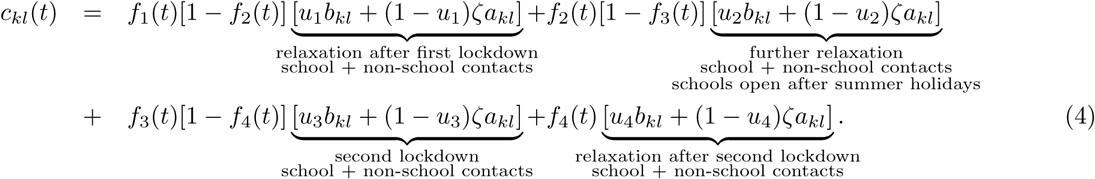

The contact rate for Scenario 2, where schools remained closed instead of reopening after the 2020 summer holidays, was modeled as

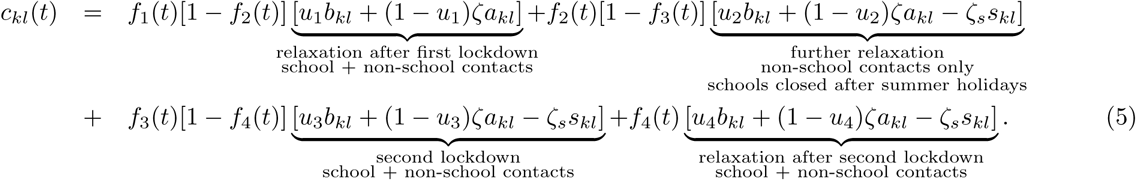

In Eq.(5), we assumed that (i) after the 2020 summer holidays school contacts are described by *ζ_s_s_kl_*, where *ζ_s_* is a mitigation factor in transmission-relevant contacts due to the protective measures in schools; consequently, *ζ_s_* was calibrated to ensure that non-school contacts did not fall below their level prior to the second relaxation if schools stay closed; (iii) school measures were similar throughout the entire period of Scenario 2; and (iii) the two-week school closure for winter holidays from 21 December 2020 till 3 January 2021 [34] had little impact on hospitalizations and, therefore, was not explicitly modeled. Results with this assumption relaxed are presented in the sensitivity analyses section below. Time-varying contact patterns for the baseline versus Scenario 1 and the baseline versus Scenario 2 are shown in Supplementary Figures S2-S4.

### Parameter estimation

The model parameters were estimated by fitting the model to the seroprevalence and hospitalization data in a Bayesian framework, as detailed in [23]. Specifically, we used the probabilistic programming language Stan [35] to sample sets of parameter vectors from the posterior distribution of the model. The modeling framework consists of three main components: (i) the prior distributions for all parameters, (ii) the dynamical model that generates epidemiological trajectories for a given parameter vector, and (iii) the observation (or error) model that links these trajectories to time series of observed data.

#### Prior distributions

For parameters *ɛ*, *θ*, *ϕ*, *ν_k_*, and *u_i_* we assigned vague or flat priors, meaning their posterior distributions were mostly inferred from the data. The duration of the exposed (*α^−^*^1^) and infectious periods (*γ^−^*^1^) had Gamma prior distributions, with 99% of the prior probability mass ranging from 2 to 5 days [36, 37] and 2.9 to 5.2 days [38], respectively. The relative susceptibilities of the 0-20 years age group, *β*_1_*_,_*_2_*_,_*_3_, and of the 20-60 years age group, *β*_4_*_,_*_5_*_,_*_6_*_,_*_7_, had log-normal priors with medians 0.23 and 0.64, respectively, and a scale parameter of 0.5 [39]. The relative susceptibility of the 60+ age group was set to *β*_8_*_,_*_9_*_,_*_10_ ≡ 1. The transmission mitigation parameter *ζ* had a N(1, 0.1) prior reflecting the belief that reduced contacts (encoded by the contact matrix *a_kl_*) were the primary determinant for the reduction in transmission during the first lockdown. The mid-point times of transition *t*_0_, *t*_2_, *t*_3_, and *t*_4_, corresponding to policy changes, were assigned normal priors with means 22, 203, 254, and 304 days post 26 February 2020, and standard deviations of 7 days. The rates *K_i_*, governing the speed at which policy changes took effect, were given Exp(1) priors, implying that the uptake of measures took around 6 days. Finally, the time point *t*_1_, marking the relaxation of the first lockdown, was assigned a prior distribution N(69 + 2.94*/K*_1_, 7), ensuring that by 4 May 2020 the effect of the relaxation reached 5% of full uptake.

#### Dynamical and observation model

Next, we solved the initial value problem defined by Eq. (1) using the Adams-Moulton method. We recorded the state of the system at each observation time *i* (*i* = 1*, … , t*_data_), where *t*_data_ corresponds to the final date in the hospitalization dataset (15 January 2021). The model was fitted to hospitalization incidence data stratified by age group. Let *h_k,i_* denote the number of hospitalizations observed in the interval [*i* − 1*, i*) for age group *k*. The cumulative incidence of hospitalizations in this time interval was approximated as *µ_k,i_* ≡ *ν_k_I_k_*(*i*). We then used a negative binomial probability mass function to model the likelihood of the observed data, given the model prediction

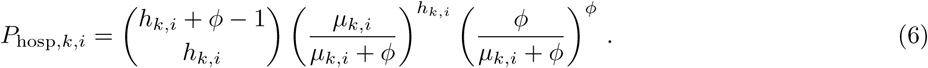

The negative binomial distribution was chosen because it allows for a larger dispersion around the mean hospitalizations compared to, for example, a Poisson distribution, allowing the model to capture large fluctuations in the hospitalization data. Relying solely on hospitalization data would have led to poor identifiability of the parameters, particularly for the hospitalization rates *ν_k_*. Therefore, we also incorporated seroprevalence data, which was sampled around day *t*_sero_ = 93 post 26 February 2020. We neglected the waning of immunity and assumed the probability of a positive serological test for a randomly selected individual in age group *k* to be *p_k_* ≡ (1 − *S_k_*(*t*_sero_)*/N_k_*). The seroprevalence data was aggregated into five age groups: [1, 10), [10, 20), [20, 40), [40, 60) and 60+ years old, as available from the data [30]. To obtain the expected fraction *q_k_* of seropositive samples in each of these age groups, we used relative population sizes. Let *L_k_* denote the number of individuals tested in age group *k* and let *ℓ_k_* represent the number of positive tests. The likelihood of observing *ℓ_k_* positive tests was modeled using a binomial distribution

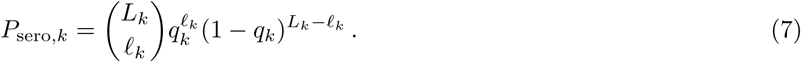

The total log-likelihood of the hospitalization and serological data was then given by

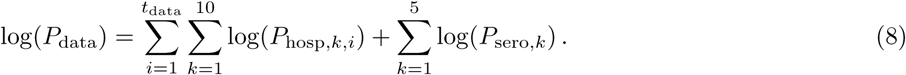

We used four parallel chains of length 1500 with a warm-up phase of length 1000, resulting in 2000 parameter samples from the posterior distribution. Estimates of the model parameters are provided in Supplementary Table S1 and Supplementary Figures S5-S7. The model fit to the hospitalizations and seroprevalence data is shown in Figure 2 and Supplementary Figures S8-S10, respectively.

**Figure 2.**
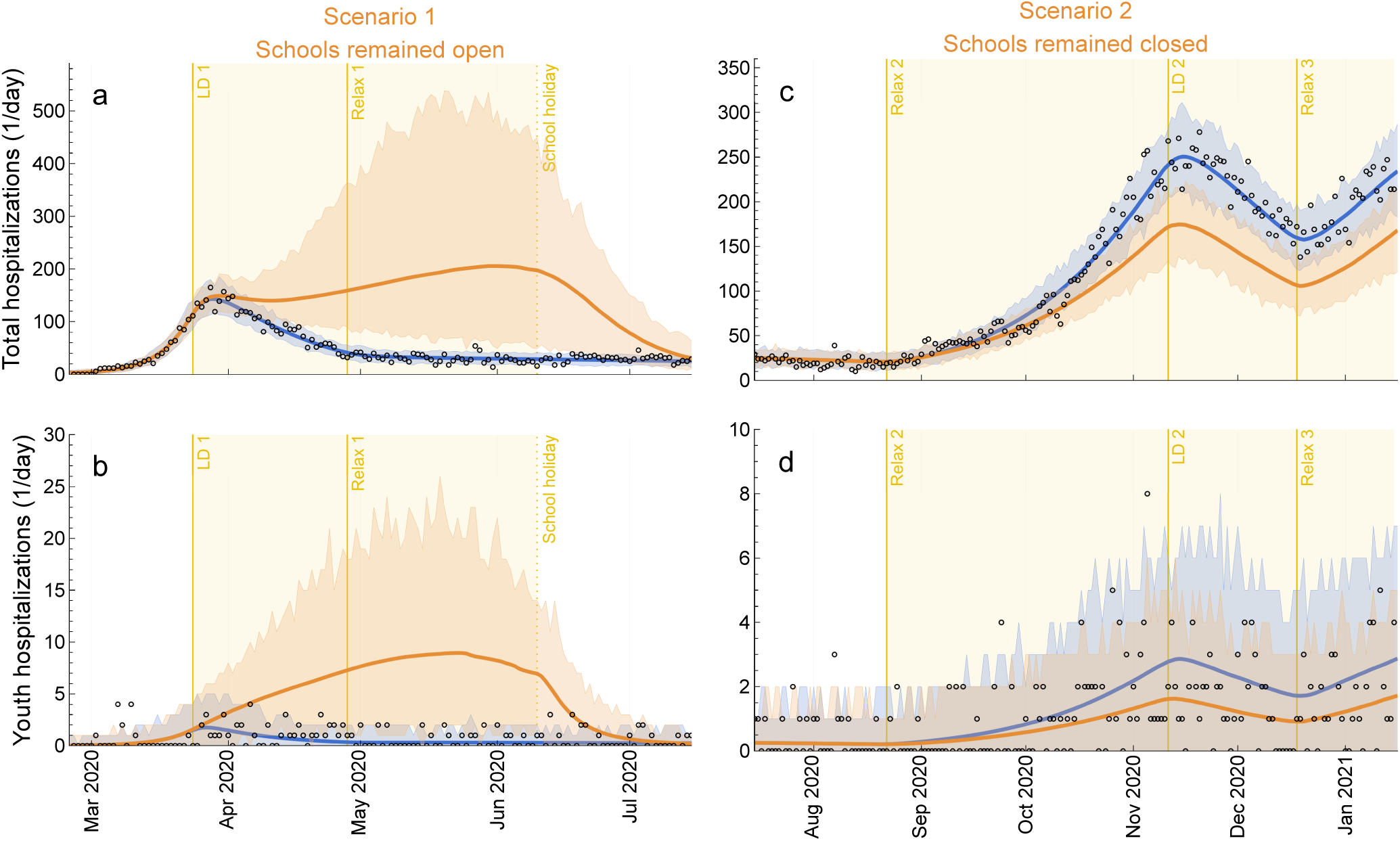
Impact of scenarios on hospitalizations. (**a**, **c**) Total and (**b**, **d**) youth (0-20 years) daily hospital admissions with COVID-19. (**a**, **b**) Scenario 1: schools remained open instead of closing during the first lockdown in spring 2020. (**c**, **d**) Scenario 2: Schools remained closed instead of reopening after the 2020 summer holidays. The circles denote daily hospital admission data. The blue lines are the median trajectories estimated from the model reflecting the baseline or real-world scenario and the blue-shaded regions correspond to the 95% CrI of the posterior predictive distribution. The orange lines and the orange-shaded regions are the respective quantities for counterfactual Scenarios 1 and 2. The yellow-shaded area represents the period when schools were open (**a**, **b**) and closed (**c**, **d**) in the counterfactual scenario analyses 1 and 2, respectively. LD: lockdown. Relax: relaxation of measures.

### Reproduction number and elasticity analysis

We calculated the time-varying effective reproduction number, *R*(*t*), which accounts for both the susceptible population and time-dependent contact rates. A value of *R*(*t*) *>* 1 indicates exponential growth in infections, whereas *R*(*t*) *<* 1 indicates a decline. *R*(*t*) was computed as the spectral radius (i.e., the largest eigenvalue) of the next-generation matrix, *K_kl_*(*t*) [40]. *K_kl_*(*t*) represents the average number of secondary infections in age group *k* caused by a single infectious individual in age group *l* at time *t* and is given by

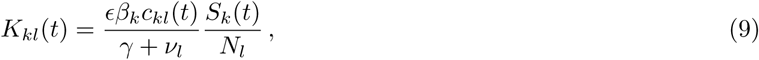

where *k, l* = 1*, … , n* and *S_k_*(*t*) is given by the solution of Eq. (1).

*R*(*t*) quantifies the transmission potential at a given time. Elasticity analysis provides insight into the drivers of changes in *R*(*t*) [41, 42]. The elasticity matrix, E*_kl_*(*t*), was defined as follows

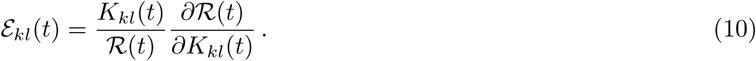

It can be shown that the sum of all elements of E*_kl_*(*t*) equals 1, i.e., 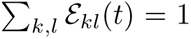[43], thus each element E*_kl_*(*t*) represents the relative contribution of *K_kl_*(*t*) to *R*(*t*). It can be further shown that the row and column sums of E*_kl_*(*t*) are equal, i.e., 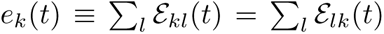 [44], indicating that the cumulative elasticities, *e_k_*(*t*), are attached only to the age group *k*, where *k* = 1*, … , n*. These quantities can therefore be interpreted as the relative contribution of each age group to *R*(*t*). Following [41], we calculated age-specific cumulative elasticities *e_k_*(*t*) as follows

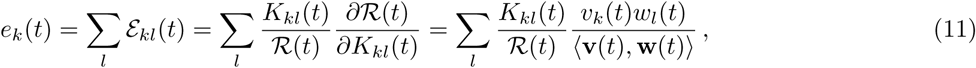

where **v**(*t*) and **w**(*t*) are the left and right eigenvectors of *K_kl_*(*t*) associated with the largest eigenvalue *R*(*t*), and ⟨·, ·⟩ denotes the dot product.

### Model outcome measures

The model outcome measures included total and age-specific daily and cumulative hospital admissions and seroprevalence, total and age-specific daily contacts, time-dependent effective reproduction number, and age-specific cumulative elasticities. For clarity, age-specific outcomes in the main text were aggregated into youth (0-20 years) and adults (20+ years) to align with key transmission patterns and policy considerations. Supplementary Figures S3-S4 and S9-S14 show results with age stratification into ten age groups. Parameter estimates were reported as medians with 95% credible intervals (CrI), based on 2000 samples from the joint posterior parameter distribution. Model projections were reported as medians with 95% CrI of the posterior predictive distribution. For computational efficiency, plots with model projections were done for 100 samples from the posterior distribution.

### Sensitivity analyses

In the sensitivity analyses, we explored the impact of the following modifications to Scenarios 1 and 2: (i) implementing the relaxation of measures in late May 2020 despite a large wave of hospitalizations in Scenario 1 (Scenario 1.1); (ii) an alternative formulation of Scenario 1 in which additional protective measures in schools reduced pre-pandemic transmission-relevant school contacts (Scenario 1.2); (iii) including school winter holidays in Scenario 2 (Scenario 2.1); (iv) an alternative formulation of contact patterns in Scenario 2 (Scenario 2.2); and (v) implementing Scenario 1 while maintaining the other measures until the end of the study period (Scenario 2.3).

## Results

### Epidemiological impact

#### Scenario 1 versus baseline

The impact of Scenarios 1 and 2 on COVID-19 hospitalizations is shown in Figure 2. For clarity, we reiterate that Scenario 1 covered the period from 26 February to 15 July 2020, assuming that schools remained open until the summer holidays began on 10 June 2020 and that the first lockdown, which targeted non-school contacts, was not relaxed in this timeframe. With school-related contacts maintained at pre-pandemic levels, this scenario resulted in a hospitalization wave substantially larger than the first pandemic wave (Figure 2 **a** and **b**). Median total hospital admissions continued to rise even after the lockdown affecting non-school contacts was implemented, only declining to levels observed in reality once schools closed for holidays.

The total cumulative hospitalizations increased to 18,028 (95% CrI 9,213–33,135) compared to 6,452 (95% CrI 6,175–6,720) at baseline, with just 3.24% (95% CrI 2.43%–3.84%) of total hospitalizations attributed to youth (0-20 years) (Supplementary Figure S13). As expected, see, e.g., [8, 23], the largest absolute number of hospitalizations among adults occurred in the oldest age groups. However, in relative terms, youth experienced the largest increase in hospitalizations compared to baseline, despite their susceptibility being approximately half that of adults over 20 years (Supplementary Table S1). Specifically, hospitalizations increased by 287% in children aged 0–5 years, 625% in those aged 5–10 years, and 707% in those aged 10–20 years. Among adults, the increase was more pronounced in younger age groups, with hospitalizations rising by 264% in those aged 20–30 years compared to 149% in those aged 80 years and older. This pattern was primarily driven by increased school-related contacts between youth and younger adults in Scenario 1 (Supplementary Figures S2-S3).

By 15 July 2020, total seroprevalence, reflecting the overall SARS-CoV-2 infection burden, increased to 15.88% (95% CrI 7.36%–28.32%) compared to 4.40% (95% CrI 3.34%–5.35%) at baseline (Supplementary Figures S15-S16). In Scenario 1, seroprevalence reached 23.01% in youth and 14.25% in adults, whereas at baseline, it was 2.52% in youth and 4.76% in adults.

#### Scenario 2 versus baseline

Scenario 2 covered the period from 15 July 2020 to 15 January 2021, assuming that, following the summer vacation, schools remained closed for in-person education throughout this timeframe. While this scenario mitigated the wave of hospitalizations that began in early autumn 2020, it did not prevent it (Figure 2 **c** and **d**).

The total cumulative hospital admissions decreased from 21,507 (95% CrI 20,985–22,105) at baseline to 15,793 (95% CrI 13,918–17,500), with youth accounting for only 0.93% (95% CrI 0.72%–1.10%) of total hospitalizations (Supplementary Figure S14). Among youth, hospitalizations declined the most in older age groups, decreasing by 35% in those aged 10–20 years, 29% in those aged 5–10 years, and 19% in those aged 0–5 years. Among adults, the reduction in hospitalizations was more pronounced in younger age groups, with a 27% decrease among those aged 20–30 years. However, differences in reductions of hospitalizations across adult age groups were generally modest. These trends were primarily driven by fewer school-related contacts among youth and younger adults following school closures (Supplementary Figures S2 and S4).

By 15 January 2021, total seroprevalence was somewhat lower than at baseline—14.82% (95% CrI 10.97%–18.52%) compared to 19.57% (95% CrI 14.93%–23.49%). Seroprevalence reached 8.14% in youth and 16.22% in adults, whereas at baseline, it was 11.87% in youth and 20.75% in adults (Supplementary Figures S15-S16).

#### Impact of school closures: Scenario 1 versus Scenario 2

Notably, the effect of school closures during the second wave in Scenario 2 was smaller than during the first lockdown in Scenario 1. This contrast is largely explained by the baseline conditions: during the second wave, transmission-relevant school contacts were already reduced by approximately 80% relative to pre-pandemic levels (Supplementary Table S1). This substantial reduction reflects the implementation of within-school mitigation measures, such as mask-wearing, hand hygiene, physical distancing, and isolation upon symptom onset, that were introduced in real-world school settings during the second half of 2020. Therefore, the incremental impact of fully closing schools in Scenario 2 was smaller, because transmission within schools was already partially suppressed at baseline. In contrast, the large effect of school closures during the first lockdown reflects the assumption in the counterfactual Scenario 1, which modeled schools remaining open throughout the lockdown with pre-pandemic levels of contact. This assumption is justified by the abrupt onset of the pandemic in early 2020, which likely left insufficient time to implement effective within-school measures. As a result, the marginal benefit of closing schools was greater during this period. We confirmed this mechanism in sensitivity analyses, which tested the effect of applying a mitigation scaling factor to school contacts in Scenario 1. Introducing a reduction in school-based contacts attenuated the benefit of school closures, supporting the interpretation above.

### Reproduction number and elasticities

To further explore the observed transmission dynamics, we analyzed the time-varying effective reproduction number, *R*(*t*), and the contributions of different age groups to *R*(*t*), measured using age-specific cumulative elasticities (Figure 3). Note that Figure 3 shows only the cumulative elasticity for youth, *e*_[0_*_,_*_20)_(*t*), as the corresponding value for adults is given by *e*_20+_(*t*) = 1 − *e*_[0_*_,_*_20)_(*t*).

**Figure 3.**
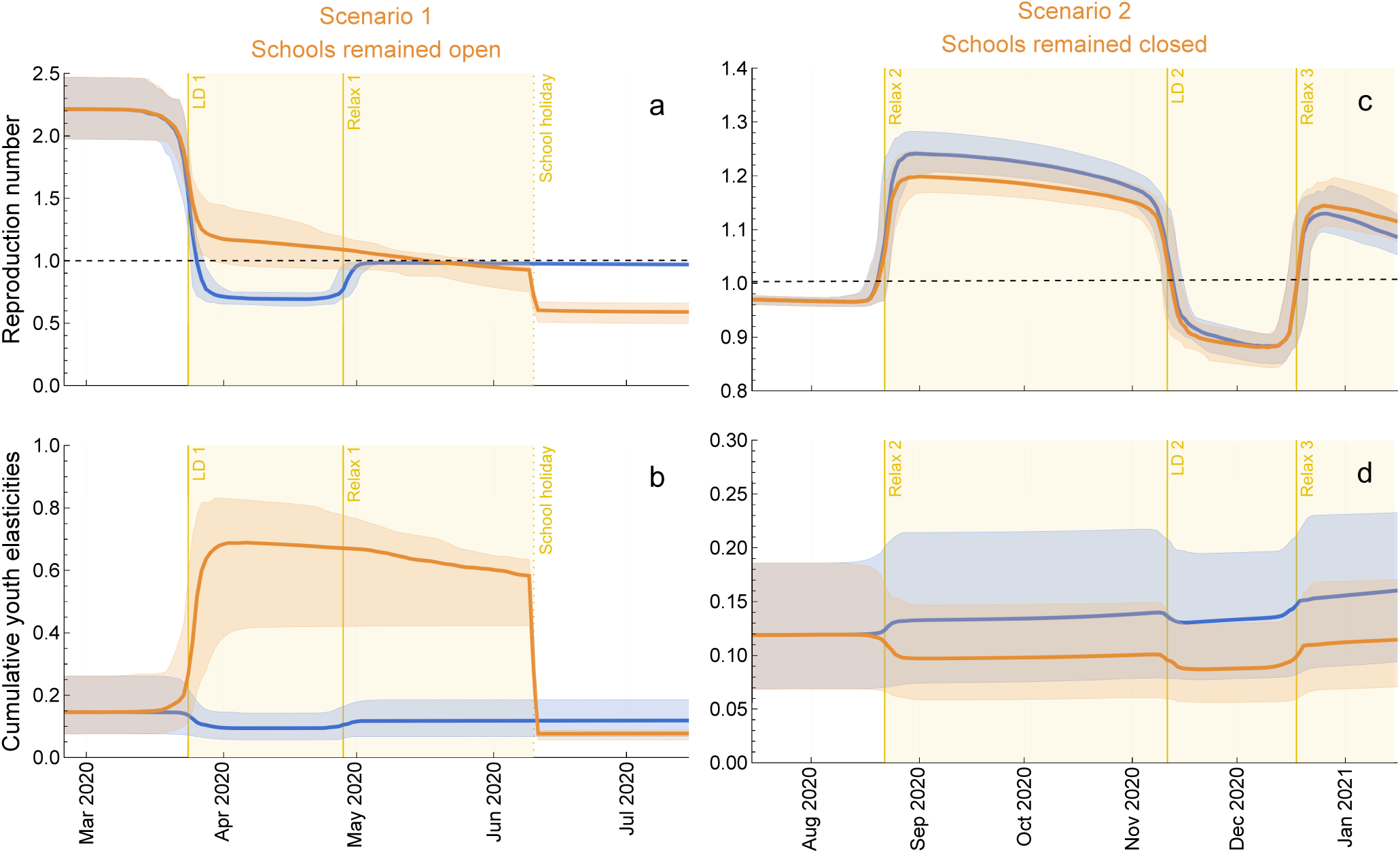
Impact of scenarios on the effective reproduction number, *R*(*t*), and relative contribution of youth to *R*(*t*). (**a**, **c**) *R*(*t*) in Scenario 1: schools remained open instead of closing during the first lockdown in spring 2020 and Scenario 2: schools remained closed instead of reopening after the 2020 summer holidays, respectively. The dashed lines correspond to *R*(*t*) = 1. (**b**, **d**) Cumulative elasticity for youth (0-20 years), *e*_[0_*_,_*_20)_(*t*), in Scenarios 1 and 2, which can be interpreted as the relative contribution of youth to *R*(*t*). Note that the cumulative elasticity for adults (20+ years) is *e*_20+_(*t*) = 1 − *e*_[0_*_,_*_20)_(*t*). The blue lines are the median trajectories estimated from the model reflecting the baseline or real-world scenario and the blue-shaded regions correspond to the 95% CrI of the posterior predictive distribution. The orange lines and the orange-shaded regions are the respective quantities for counterfactual Scenarios 1 and 2. The yellow-shaded area represents the period when schools were open (**a**, **b**) and closed (**c**, **d**) in Scenarios 1 and 2, respectively. LD: lockdown. Relax: relaxation of measures.

#### Scenario 1 versus baseline

In Scenario 1, where schools remained open, *R*(*t*) decreased rapidly but stayed above 1 despite the lockdown targeting non-school contacts (Figure 3 **a**). It then gradually declined as the susceptible population depleted, crossing 1 on 17 May 2020 when the pandemic wave peaked (Figure 2 **a**). With the start of the school holidays on 10 June 2020, *R*(*t*) dropped sharply to 0.72 (95% CrI 0.60–0.78), which is slightly below the baseline estimate during the first lockdown (0.69, 95% CrI 0.64–0.75). Thereafter, it remained stable, as the lockdown was not lifted in Scenario 1, but the susceptible population was lower than in reality. The trajectory of *R*(*t*) was reflected in the contribution of youth to overall transmission (Figure 3 **b**). Their contribution increased rapidly from 0.16 (95% CrI 0.08–0.28) to 0.69 (95% CrI 0.40–0.83) following the lockdown, then slowly declined to 0.58 (95% CrI 0.42–0.64) as the depletion of the susceptible population accelerated while schools remained open, before dropping abruptly to 0.08 (95% CrI 0.06–0.09) at the start of the school holidays.

#### Scenario 2 versus baseline

In Scenario 2, keeping schools closed during the second relaxation of non-school measures led to only a slight reduction in *R*(*t*), which remained well above 1 (Figure 3 **c**). As a result, this scenario mitigated but did not prevent the autumn wave (Figure 2 **c**). Notably, *R*(*t*) remained below the baseline value for most of the period. However, during the third relaxation of measures, it exceeded the baseline value due to a larger susceptible population in Scenario 2 compared to reality. Throughout Scenario 2, the relative contribution of youth to *R*(*t*) remained only slightly lower than at baseline, ranging between 0.09 (95% CrI 0.06–0.13) and 0.11 (95% CrI 0.07–0.17). The gradual increase in the cumulative elasticity for youth can be attributed to their share of the susceptible population, as all other relevant estimated parameters remain either constant throughout the entire analysis period or unchanged within distinct regimes of this period (Supplementary Figures S2 and S4).

#### Uncertainty in projections: Scenario 1 versus Scenario 2

The larger uncertainty in projections for Scenario 1 compared to Scenario 2 reflects the fact that *R*(*t*) was close to 1 for some of the simulated trajectories. In this regime, the system is highly sensitive to small differences in parameters, resulting in greater variability in the model outcomes (Supplementary Figure S17). This contrasts with Scenario 2, where *R*(*t*) remained well above 1 for all trajectories, leading to more predictable epidemic behavior. This observation highlights not only the substantial impact of school closures during the first lockdown, but also the greater uncertainty associated with counterfactual projections in that period.

### Robustness of results

The transmission dynamics presented in the main text were qualitatively robust to alternative formulations of Scenarios 1 and 2. Results for Scenario 1, in which the first lockdown was relaxed despite rising hospitalizations, are shown in Supplementary Figure S18 (Scenario 1.1). Supplementary Figure S19 shows a variation of Scenario 1 incorporating additional measures to reduce pre-pandemic transmission-relevant contacts in schools during the period when schools remained open (Scenario 1.2). In these two analyses, the first wave of hospitalizations was substantially larger and smaller, respectively, than in the main results, yet in both cases it exceeded the first wave observed in the data.

For Scenario 2, the two-week school closure during the winter holidays around Christmas and New Year 2021 had a minor impact on hospitalizations (Scenario 2.1; Supplementary Figure S20). A second variant of Scenario 2, which used an alternative formulation of contact patterns, led to a smaller hospitalization wave compared to our original Scenario 2 (Scenario 2.2; Supplementary Figure S21). Overall, both sensitivity analyses for Scenario 2 support the robustness of our findings, i.e., school closures after the summer holidays might have reduced the magnitude of the wave, but they would not have prevented it altogether.

## Discussion

Our findings demonstrate that school closures can be an important component of the pandemic response, but their effectiveness is highly context-dependent. During the first wave of the pandemic, when no mitigation measures were in place within schools, keeping schools open would likely have sustained transmission and prevented *R*(*t*) from falling below 1, despite broader lockdown efforts. In this setting, school closures contributed substantially to reducing transmission and were essential for achieving control of virus spread.

In contrast, the modeled effect of school closures during the autumn of 2020 was markedly smaller. By this time, various mitigation measures had already reduced transmission within schools, while transmission outside school settings remained elevated due to the easing of restrictions affecting non-school contacts. In this context, closing schools alone did not suffice to bring *R*(*t*) below 1 and failed to contain the second pandemic wave. This distinction underscores a key insight, also highlighted by other studies for the Netherlands and the UK [8, 14, 45]: the epidemiological impact of school closures depends not only on transmission dynamics within schools but also on the strength of control measures in the general population.

From a policymaker’s perspective, decisions about school closures must balance their epidemiological impact with broader societal consequences [46, 47]. While school-based transmission contributed to the overall spread of the virus, in agreement with other studies we found that youth typically experienced lower susceptibility [8, 23, 48–50] and their direct contribution to hospitalizations was limited [8, 23, 51–53]. Thus, the primary justification for school closures lies in their potential to reduce transmission in the wider community, thereby preventing a surge of hospitalizations. However, if closures fail to bring *R*(*t*) below 1 and do not meaningfully alter key epidemiological outcomes, their benefits may be outweighed by societal costs such as learning losses and deterioration in mental health among youth [46, 47]. Future modeling studies should explicitly evaluate these trade-offs, quantifying both the expected epidemiological benefits and the broader societal costs of school closures.

To our knowledge, this is the first study to retrospectively assess the impact of school openings and closures on SARS-CoV-2 transmission during the pre-vaccination period in Portugal. Most prior transmission modeling studies either did not explicitly incorporate school interventions [22,25] or focused on forward projections of potential school closures later in the pandemic [6]. Unlike prior studies [2, 19], our model was age-structured and formally fitted to empirical data, allowing us to estimate key age-specific parameters such as susceptibility, hospitalization rates, and contact patterns that were consistent with the observed course of the pandemic. A distinctive feature of our modeling framework is that changes in contact patterns, including their timing, speed, and age-specific mixing, were inferred directly from the data rather than imposed a priori [6, 11, 14].

The model accurately reproduced age-specific hospitalization and seroprevalence data, lending credibility to its projections. Moreover, our conclusions remained qualitatively robust across alternative formulations of school closure and reopening scenarios. While most previous modeling studies focused on estimating reproduction numbers and population-level epidemiological outcomes such as hospitalizations or infections [2, 5, 6, 22], we additionally applied elasticity analysis to quantify the relative contributions of different age groups to transmission over time. Consistent with earlier studies from Portugal and Belgium, we found that adults were the primary contributors to the reproduction number in 2020 [16, 54]. However, unlike those studies, which evaluated contributions only at the onset of the pandemic [16, 55–57] or at selected time points [54, 58], our analysis captured how these contributions evolved over time and identified the key drivers behind these shifts.

Finally, our model is parsimonious yet generalizable and could be adapted to study the transmission dynamics of other novel respiratory pathogens transmitted via close person-to-person contact, provided that comparable empirical data are available. Nevertheless, caution is needed when extrapolating our findings to other pathogens, as their epidemiological characteristics may differ substantially from those of SARS-CoV-2. For instance, during the 2009 H1N1 influenza pandemic, youth exhibited higher susceptibility and experienced more severe disease than has been observed with SARS-CoV-2 [59, 60].

While our model was formally fitted to empirical data, our study has several limitations. First, it relied on publicly available contact and seroprevalence data with limited granularity. More detailed data on interactions within and outside school settings could have reduced the uncertainty in the model projections. Additionally, seroprevalence data may be biased, as serological assays often failed to detect antibodies following asymptomatic infections [61,62], which are particularly common in youth [51]. As a result, our analysis may have underestimated the role of youth in overall transmission. Second, the model did not account for seasonality in transmission because the empirical data did not allow for disentangling seasonal effects from those of non-pharmaceutical interventions. Consequently, external factors such as climate variation and seasonal behavioral changes [63] may have influenced transmission dynamics in ways not captured by our analysis. Third, individual mitigation measures such as mask-wearing or symptom-based isolation were not modeled explicitly. Instead, we assumed that their aggregate effects were reflected in the overall reduction of school- and non-school transmission-relevant contacts. More detailed models would be better suited to evaluate the impact of individual measures within schools [64–67]. Finally, the model did not distinguish between primary and secondary school settings. Instead, school closures and openings were modeled as changes to the overall school contact matrix. Nonetheless, we stratified the youth population into three age groups (0–5, 5–10, and 10–20 years), each with age-specific parameters, consistent with the granularity of the available data, allowing us to capture age-specific transmission dynamics within these groups. The value of age-stratified transmission models in understanding the dynamics of SARS-CoV-2 has been emphasized in prior studies for the Netherlands and Germany [68, 69].

## Conclusions

In conclusion, we found that school closures played a critical role in suppressing virus spread during the initial pandemic phase in Portugal. However, in later phases, when within-school mitigation measures were in place and transmission was largely driven by non-school settings, the impact of schools diminished. These findings underscore that the epidemiological impact of school closures is highly context-dependent, influenced by mitigation measures within schools and the intensity of transmission outside them. While closures can support control efforts, especially early into a pandemic, their benefits must be weighed against substantial societal costs. Our results suggest that integrated strategies combining school-based and broader community interventions are essential to effectively reduce transmission during future pandemics.

## Data Availability

All data produced in the present study will be available upon publication.

## Supplementary information

The Supplementary Material provides additional figures, tables, and further details of this study. The data and codes reproducing the results of this study will be available upon publication at https://github.com/rsescosio/COVID19-PT-retrospective. For peer review, the contents of the GitHub repository have been provided in a compressed zip format.

## Acknowledgements

We appreciate the valuable discussions with members of the Infectious Disease Modeling Group at the University Medical Center Utrecht in the Netherlands.

## Author contributions

Conceptualization: GR; Methodology: CHvD and GR; Software: BC and RAE; Validation: CHvD, OB, MB, and AN; Formal Analysis: BC, RAE, and GR; Investigation: BC, RAE, and GR; Resources: AN and GR; Data Curation: CHvD and GR; Writing–Original Draft: BC, RAE, CHvD, and GR; Writing–Review & Editing: All authors; Visualization: RAE; Supervision: AN and GR; Project Administration: AN and GR; Funding Acquisition: GR.

## Supplementary Material

## 1 Summary of the model parameters

**Table S1.**
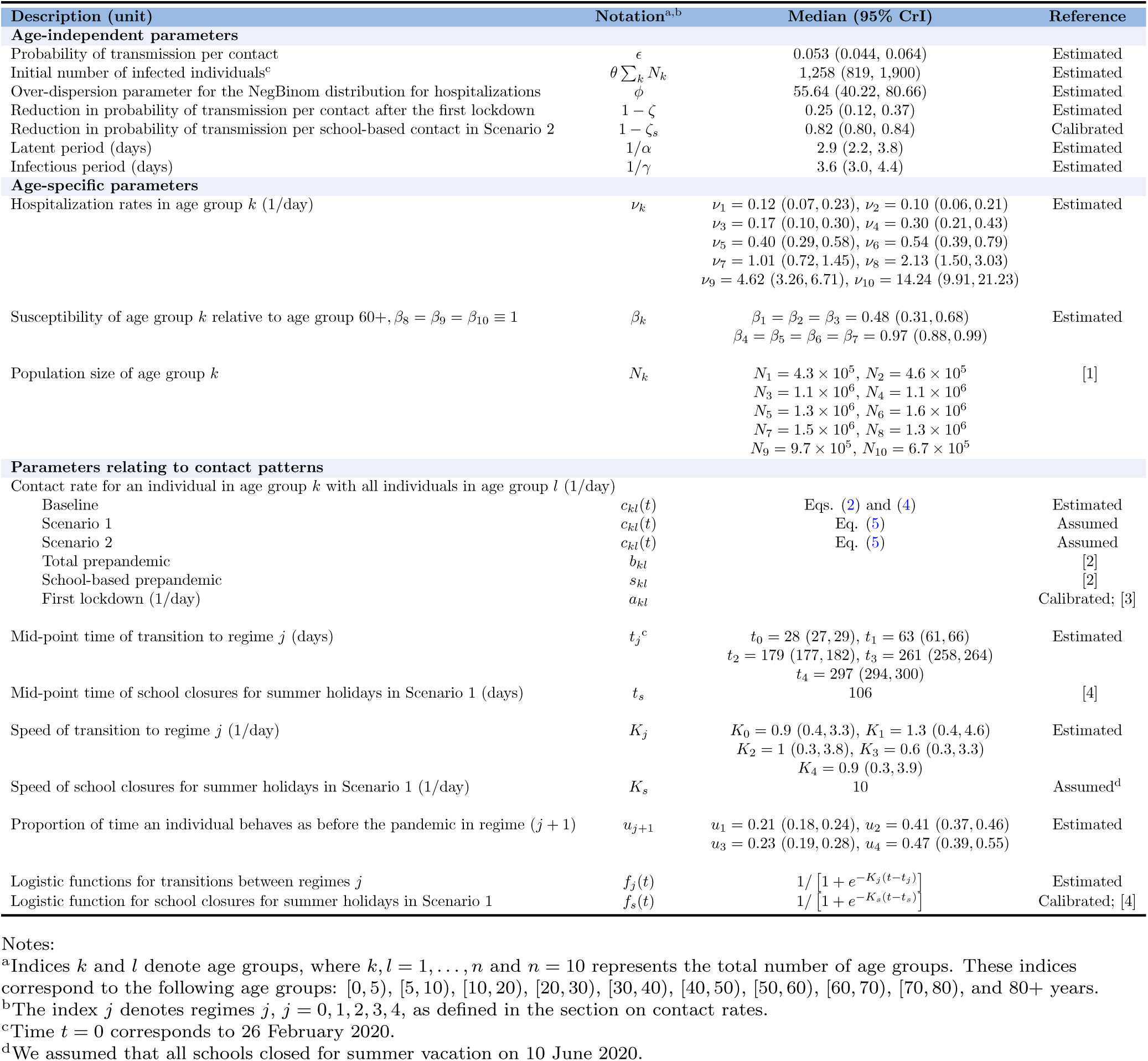
Summary of the model parameters. This table provides an overview of all model parameters as defined in various sections of the Methods.

| Description (unit) | Notation<sup>a,b</sup> | Median (95% CrI) | Reference |
| --- | --- | --- | --- |
| **Age-independent parameters** |  |  |  |
| Probability of transmission per contact | $\epsilon$ | 0.053 (0.044, 0.064) | Estimated |
| Initial number of infected individuals<sup>c</sup> | $\theta \sum_k N_k$ | 1,258 (819, 1,900) | Estimated |
| Over-dispersion parameter for the NegBinom distribution for hospitalizations | $\phi$ | 55.64 (40.22, 80.66) | Estimated |
| Reduction in probability of transmission per contact after the first lockdown | $1 - \zeta$ | 0.25 (0.12, 0.37) | Estimated |
| Reduction in probability of transmission per school-based contact in Scenario 2 | $1 - \zeta_s$ | 0.82 (0.80, 0.84) | Calibrated |
| Latent period (days) | $1/\alpha$ | 2.9 (2.2, 3.8) | Estimated |
| Infectious period (days) | $1/\gamma$ | 3.6 (3.0, 4.4) | Estimated |
| **Age-specific parameters** |  |  |  |
| Hospitalization rates in age group $k$ (1/day) | $\nu_k$ | $\nu_1 = 0.12$ (0.07, 0.23), $\nu_2 = 0.10$ (0.06, 0.21)<br>$\nu_3 = 0.17$ (0.10, 0.30), $\nu_4 = 0.30$ (0.21, 0.43)<br>$\nu_5 = 0.40$ (0.29, 0.58), $\nu_6 = 0.54$ (0.39, 0.79)<br>$\nu_7 = 1.01$ (0.72, 1.45), $\nu_8 = 2.13$ (1.50, 3.03)<br>$\nu_9 = 4.62$ (3.26, 6.71), $\nu_{10} = 14.24$ (9.91, 21.23) | Estimated |
| Susceptibility of age group $k$ relative to age group 60+, $\beta_8 = \beta_9 = \beta_{10} \equiv 1$ | $\beta_k$ | $\beta_1 = \beta_2 = \beta_3 = 0.48$ (0.31, 0.68)<br>$\beta_4 = \beta_5 = \beta_6 = \beta_7 = 0.97$ (0.88, 0.99) | Estimated |
| Population size of age group $k$ | $N_k$ | $N_1 = 4.3 \times 10^5$, $N_2 = 4.6 \times 10^5$<br>$N_3 = 1.1 \times 10^6$, $N_4 = 1.1 \times 10^6$<br>$N_5 = 1.3 \times 10^6$, $N_6 = 1.6 \times 10^6$<br>$N_7 = 1.5 \times 10^6$, $N_8 = 1.3 \times 10^6$<br>$N_9 = 9.7 \times 10^5$, $N_{10} = 6.7 \times 10^5$ | [1] |
| **Parameters relating to contact patterns** |  |  |  |
| Contact rate for an individual in age group $k$ with all individuals in age group $l$ (1/day) | | | |
| Baseline | $c_{kl}(t)$ | Eqs. (2) and (4) | Estimated |
| Scenario 1 | $c_{kl}(t)$ | Eq. (5) | Assumed |
| Scenario 2 | $c_{kl}(t)$ | Eq. (5) | Assumed |
| Total prepandemic | $b_{kl}$ | | [2] |
| School-based prepandemic | $s_{kl}$ | | [2] |
| First lockdown (1/day) | $a_{kl}$ | | Calibrated; [3] |
| Mid-point time of transition to regime $j$ (days) | $t_j^c$ | $t_0 = 28$ (27, 29), $t_1 = 63$ (61, 66)<br>$t_2 = 179$ (177, 182), $t_3 = 261$ (258, 264)<br>$t_4 = 297$ (294, 300) | Estimated |
| Mid-point time of school closures for summer holidays in Scenario 1 (days) | $t_s$ | 106 | [4] |
| Speed of transition to regime $j$ (1/day) | $K_j$ | $K_0 = 0.9$ (0.4, 3.3), $K_1 = 1.3$ (0.4, 4.6)<br>$K_2 = 1$ (0.3, 3.8), $K_3 = 0.6$ (0.3, 3.3)<br>$K_4 = 0.9$ (0.3, 3.9) | Estimated |
| Speed of school closures for summer holidays in Scenario 1 (1/day) | $K_s$ | 10 | Assumed<sup>d</sup> |
| Proportion of time an individual behaves as before the pandemic in regime $(j + 1)$ | $u_{j+1}$ | $u_1 = 0.21$ (0.18, 0.24), $u_2 = 0.41$ (0.37, 0.46)<br>$u_3 = 0.23$ (0.19, 0.28), $u_4 = 0.47$ (0.39, 0.55) | Estimated |
| Logistic functions for transitions between regimes $j$ | $f_j(t)$ | $1 / [1 + e^{-K_j(t-t_j)}]$ | Estimated |
| Logistic function for school closures for summer holidays in Scenario 1 | $f_s(t)$ | $1 / [1 + e^{-K_s(t-t_s)}]$ | Calibrated; [4] |

Notes:
<sup>a</sup>Indices $k$ and $l$ denote age groups, where $k, l = 1, \dots, n$ and $n = 10$ represents the total number of age groups. These indices correspond to the following age groups: [0, 5), [5, 10), [10, 20), [20, 30), [30, 40), [40, 50), [50, 60), [60, 70), [70, 80), and 80+ years.
<sup>b</sup>The index $j$ denotes regimes $j$ , $j = 0, 1, 2, 3, 4$ , as defined in the section on contact rates.
<sup>c</sup>Time $t = 0$ corresponds to 26 February 2020.
<sup>d</sup>We assumed that all schools closed for summer vacation on 10 June 2020.

## 2 Contact matrices

**Figure S1.**
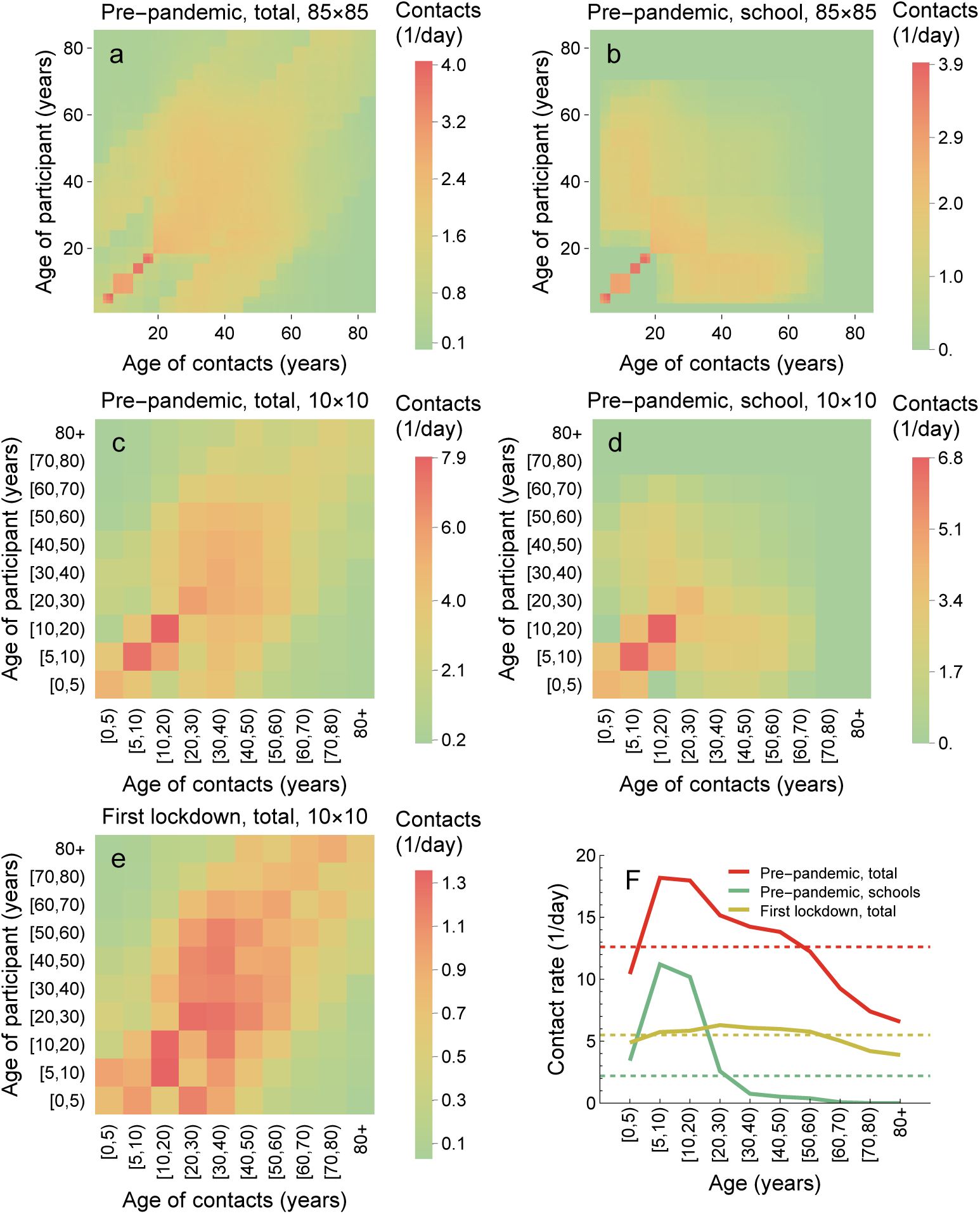
Contact matrices. The 86 × 86 pre-pandemic contact matrices for (**a**) total and (**b**) school-based contacts were taken from Mistry et al [2]. Using the Portuguese demographic data, we obtained the coarse-grained 10 × 10 pre-pandemic contact matrices for (**c**) total and (**d**) school-based contacts. Due to the lack of contact data for Portugal during the first lockdown, such as that provided by Mistry et al [2] for the pre-pandemic period, we followed our previous approach [5] and inferred the Portuguese total contact matrix for this period (**e**) using data from the Netherlands [3], where a comparable set of interventions was implemented. The average number of contacts for an individual in a given age group for the three 10 × 10 contact matrices are shown in (**f**).

## 3 Contact patterns

**Figure S2.**
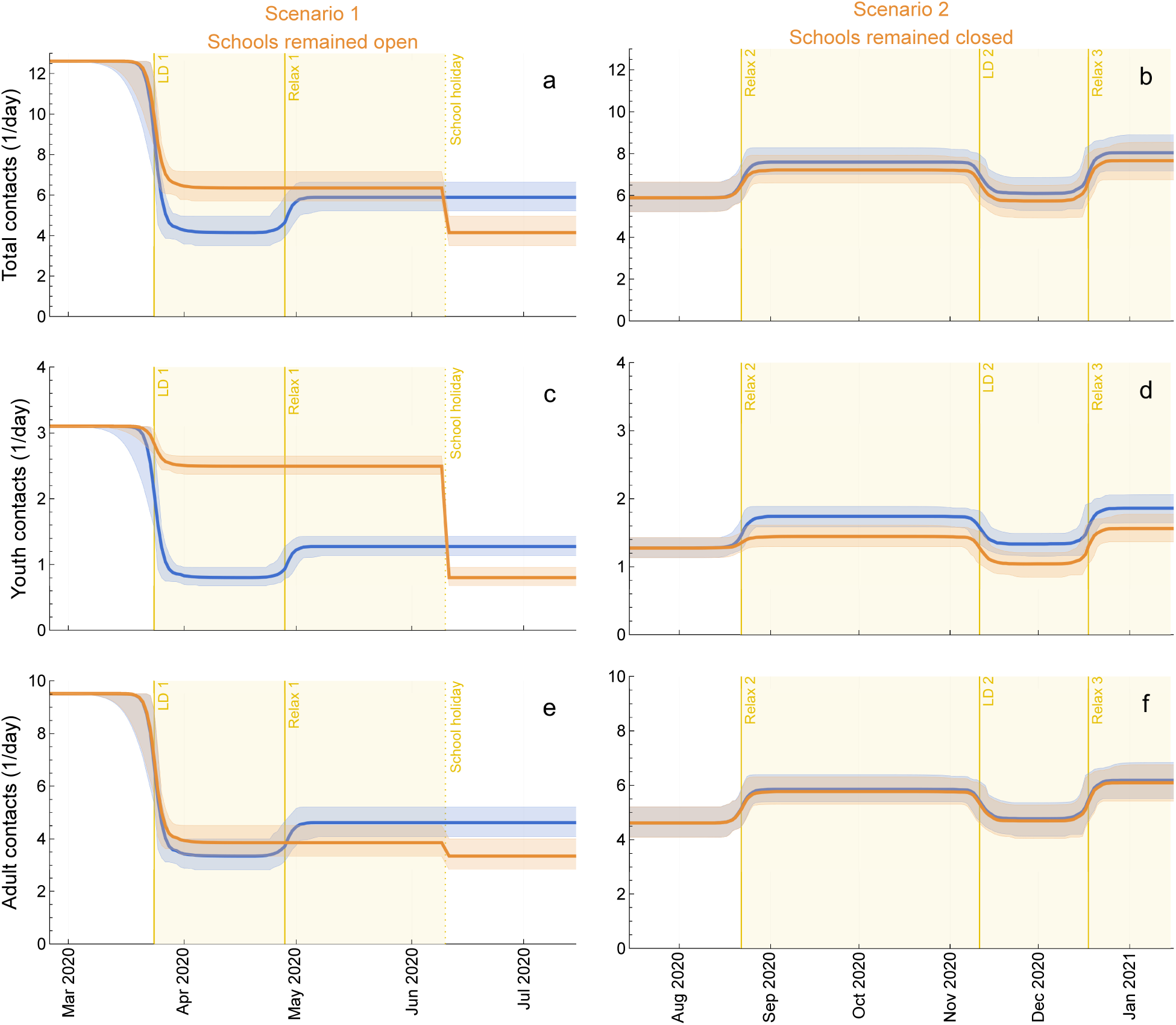
Impact of scenarios on contact rates. (**a**, **b**) Total, (**c**, **d**) youth (0-20 years), and (**e**, **f**) adult (20+ years) contact rates in (**a**, **c**, **e**) Scenario 1: schools remained open instead of closing during the first lockdown in spring 2020 and (**b**, **d**, **f**) Scenario 2: schools remained closed instead of reopening after the 2020 summer holidays. The blue lines are the median trajectories estimated from the model and the blue-shaded regions correspond to the 95% CrI of the posterior predictive distribution. The orange lines and the orange-shaded regions are the respective quantities in Scenarios 1 and 2. The yellow-shaded area represents the period when schools were open (**a**, **c**, **e**) and closed (**b**, **d**, **f**) in the counterfactual scenario analyses 1 and 2, respectively. LD: lockdown. Relax: relaxation of measures.

**Figure S3.**
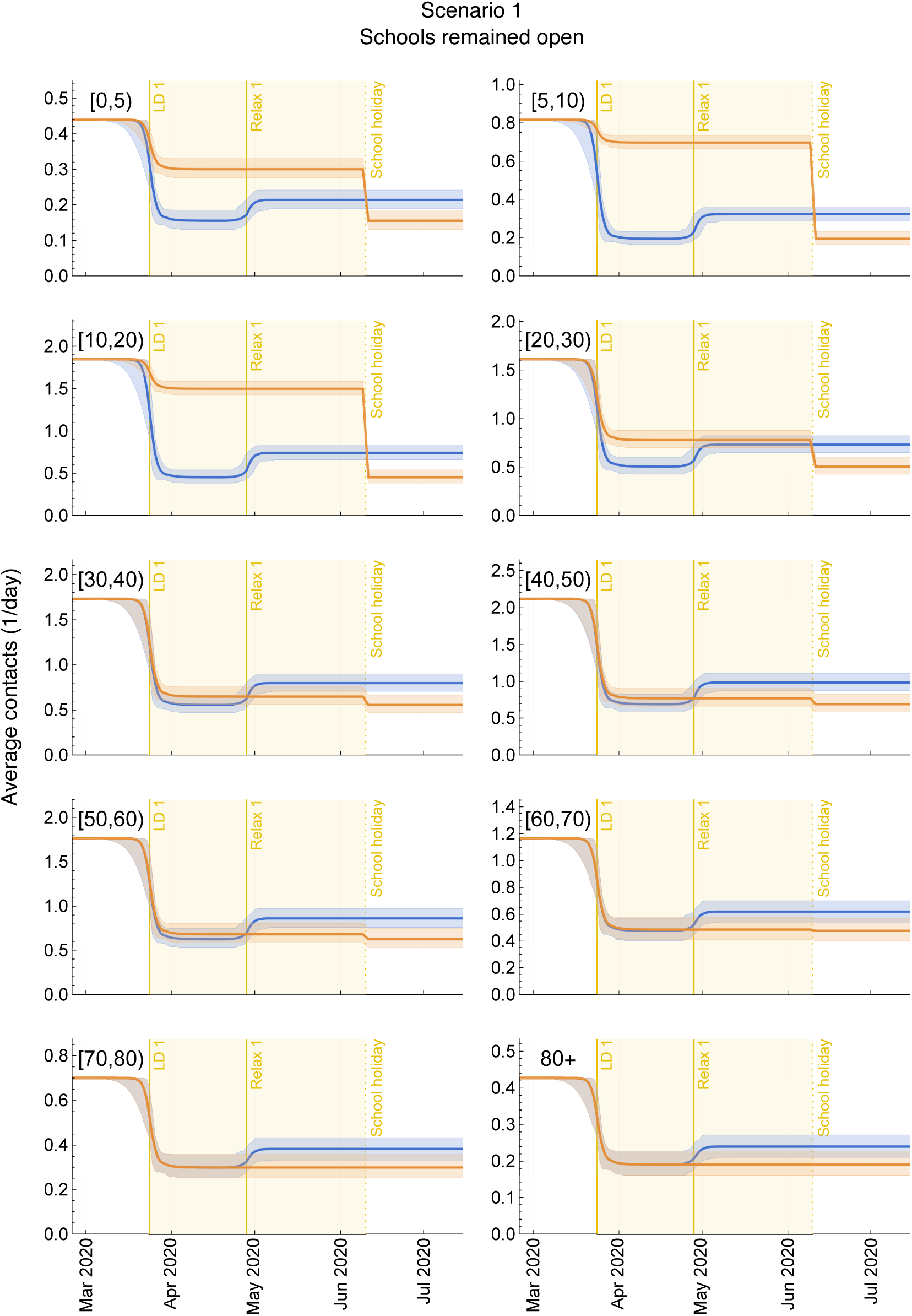
Impact of Scenario 1 on contact rates in ten age groups. Contact rates in Scenario 1: schools remained open instead of closing during the first lockdown in spring 2020 for the ten age groups, as annotated. The blue lines are the median trajectories estimated from the model and the blue-shaded regions correspond to the 95% CrI of the posterior predictive distribution. The orange lines and the orange-shaded regions are the respective quantities for Scenario 1. The yellow-shaded area represents the period when schools were open in the counterfactual scenario analyses 1. LD: lockdown. Relax: relaxation of measures.

**Figure S4.**
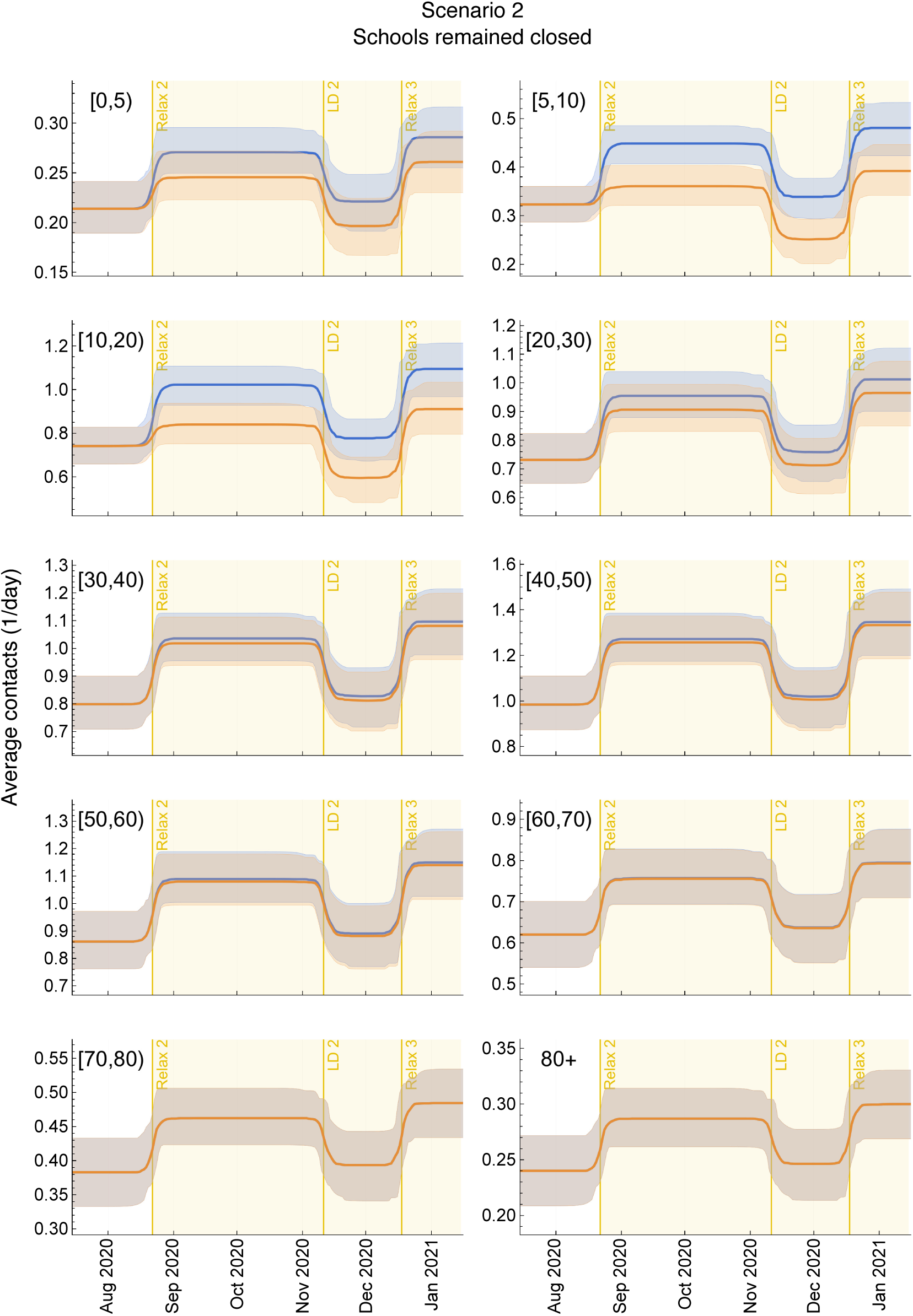
Impact of Scenario 2 on contact rates in ten age groups. Contact rates in Scenario 2: schools remained closed instead of reopening after the 2020 summer holidays for the ten age groups, as annotated. The blue lines are the median trajectories estimated from the model and the blue-shaded regions correspond to the 95% CrI of the posterior predictive distribution. The orange lines and the orange-shaded regions are the respective quantities for Scenario 2. The yellow-shaded area represents the period when schools were closed in the counterfactual scenario analyses 2. LD: lockdown. Relax: relaxation of measures.

## 4 Estimated parameters

**Figure S5.**
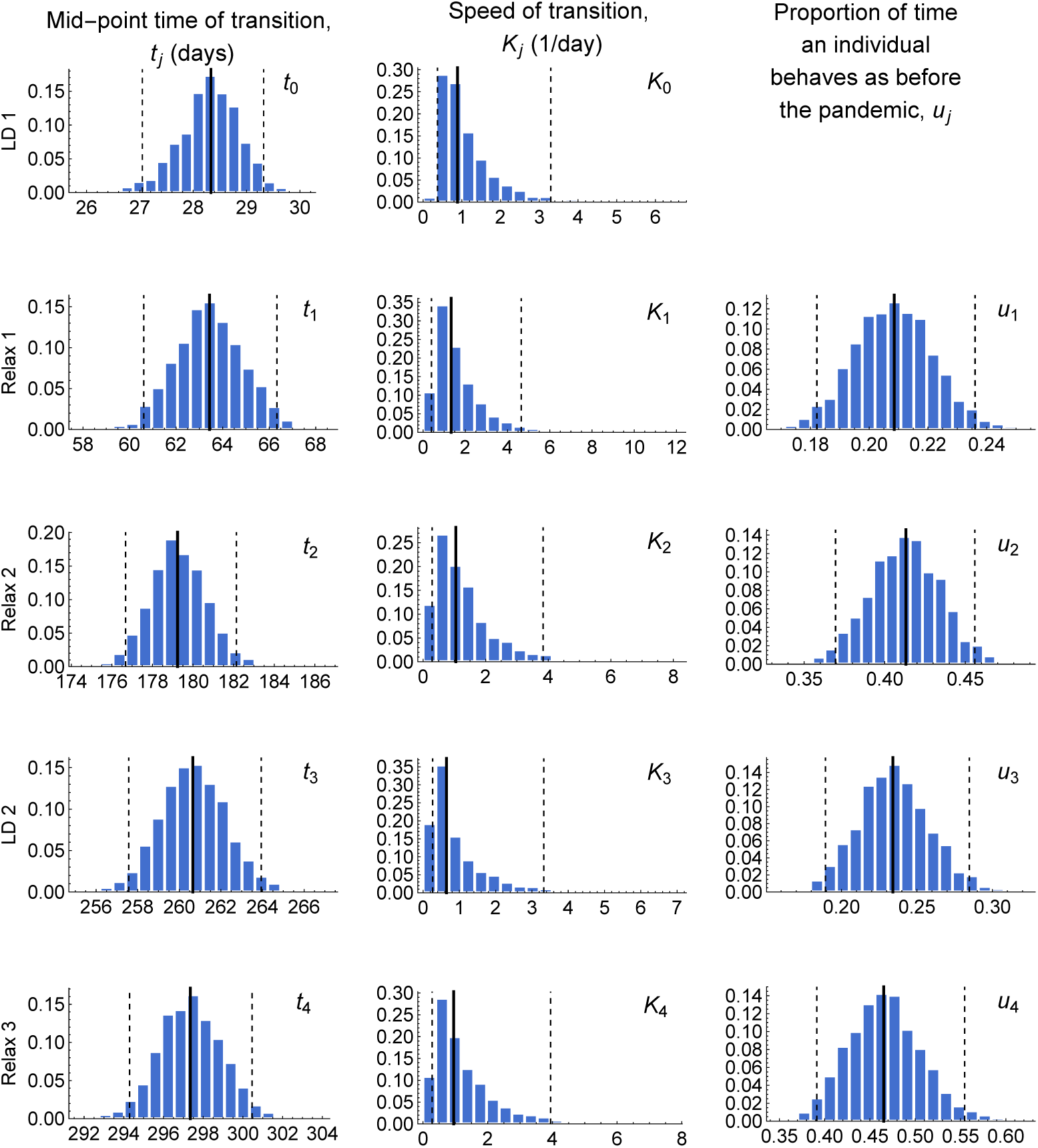
Estimated parameters describing contact patterns. The histograms are based on 2000 parameter samples from the posterior distribution for the (left) mid-point time of transition to regime *j*, *t_j_*, (center) speed of transition, *K_j_*, and (right) proportion of time an individual behaves as before the pandemic in regime (*j* + 1), *u_j_*, for each regime *j* = 0, 1, 2, 3, 4, annotated in rows. The solid and the dashed black lines correspond to the median and the 95% CrI, both of which are tabulated in Table S1. LD: lockdown. Relax: relaxation.

**Figure S6.**
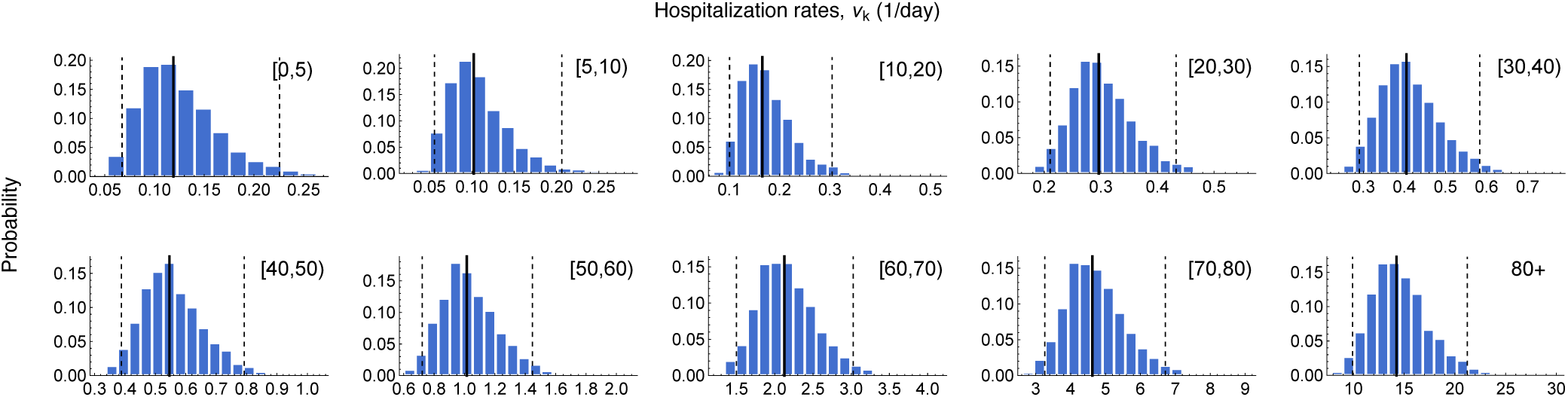
Estimated hospitalization rates. The histograms are based on 2000 parameter samples from the posterior distribution for the age-specific hospitalization rate in age-group *k*, *ν_k_*, where *k* = 1*, … ,* 10. The solid and the dashed black lines correspond to the median and the 95% CrI, both of which are tabulated in Table S1.

**Figure S7.**
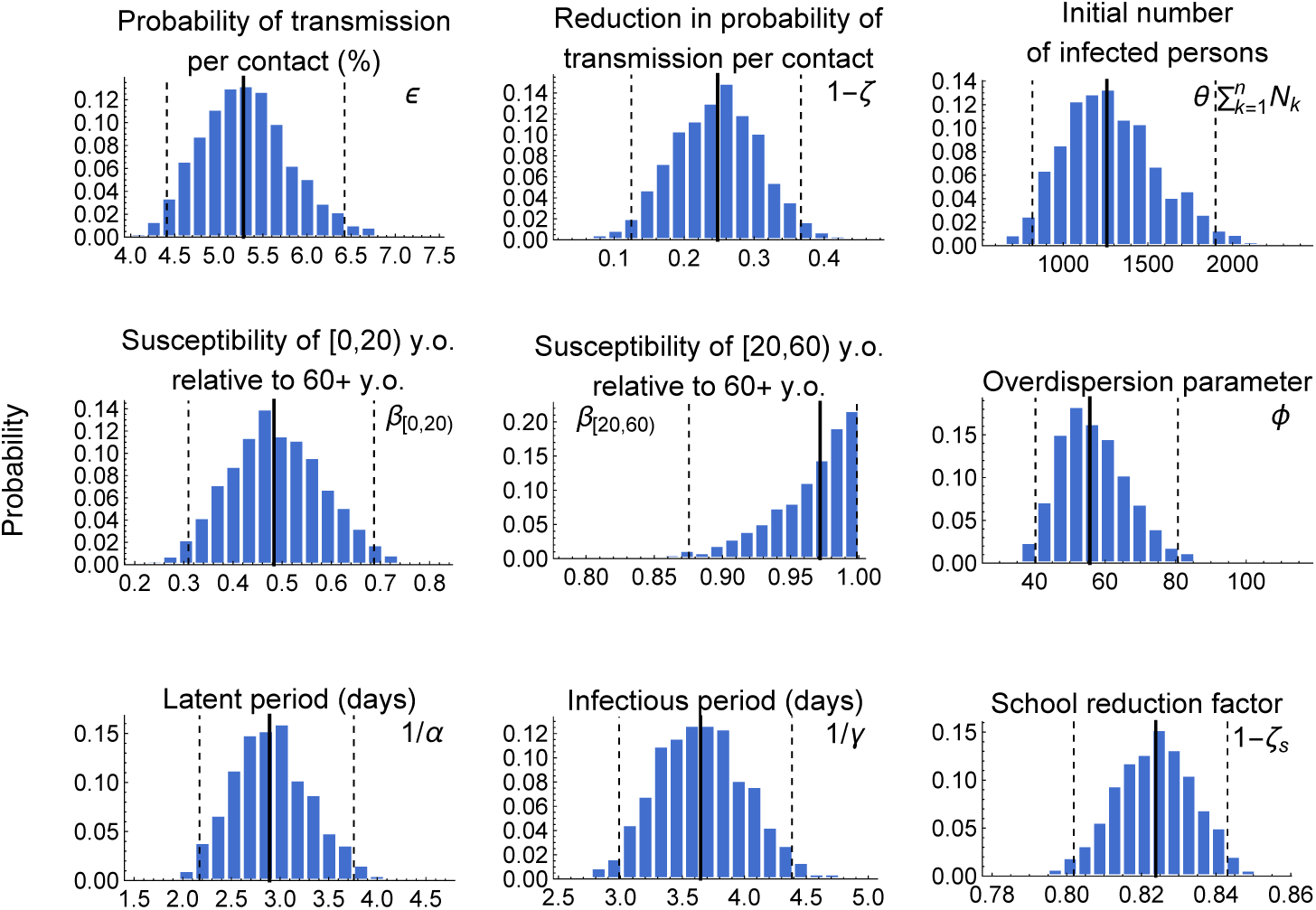
Estimates of other parameters. The histograms are based on 2000 parameter samples from the posterior distribution for the estimated model parameters, each described by their respective titles. The solid and the dashed black lines correspond to the median and the 95% CrI, both of which are listed in Table S1. All parameters were estimated using a Bayesian framework, except for *ζ_s_*, which was calibrated. For this, school contacts after the 2020 summer holidays were described by *ζ_s_s_kl_*, where *ζ_s_* is a mitigation factor in school transmission-relevant contacts due to the protective measures in schools and *s_kl_* is pre-pandemic school contact matrix. Consequently, *ζ_s_* was calibrated using linear programming to ensure that non-school contacts did not fall below their levels prior to the second relaxation if schools remain closed in Scenario 2.

## 5 Model fit to seroprevalence data

**Figure S8.**
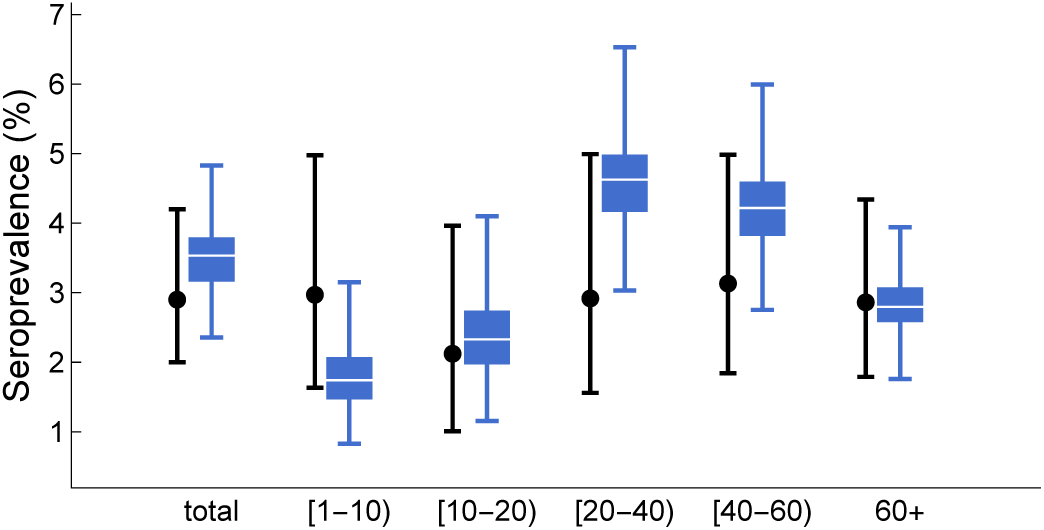
Model fit to COVID-19 seroprevalence. Both total and age-specific seroprevalence are shown in the plot. The black circles with 95% confidence intervals represent the data obtained from the first serological survey (ISNCOVID-19) conducted in Portugal between 21 May 2020 and 8 July 2020 [6] used for fitting the model. The blue box plots represent the marginal posterior distribution of the seroprevalence on 28 May 2020 based on 2000 samples from the joint posterior parameter distribution.

## 6 Model fit to hospitalization data

**Figure S9.**
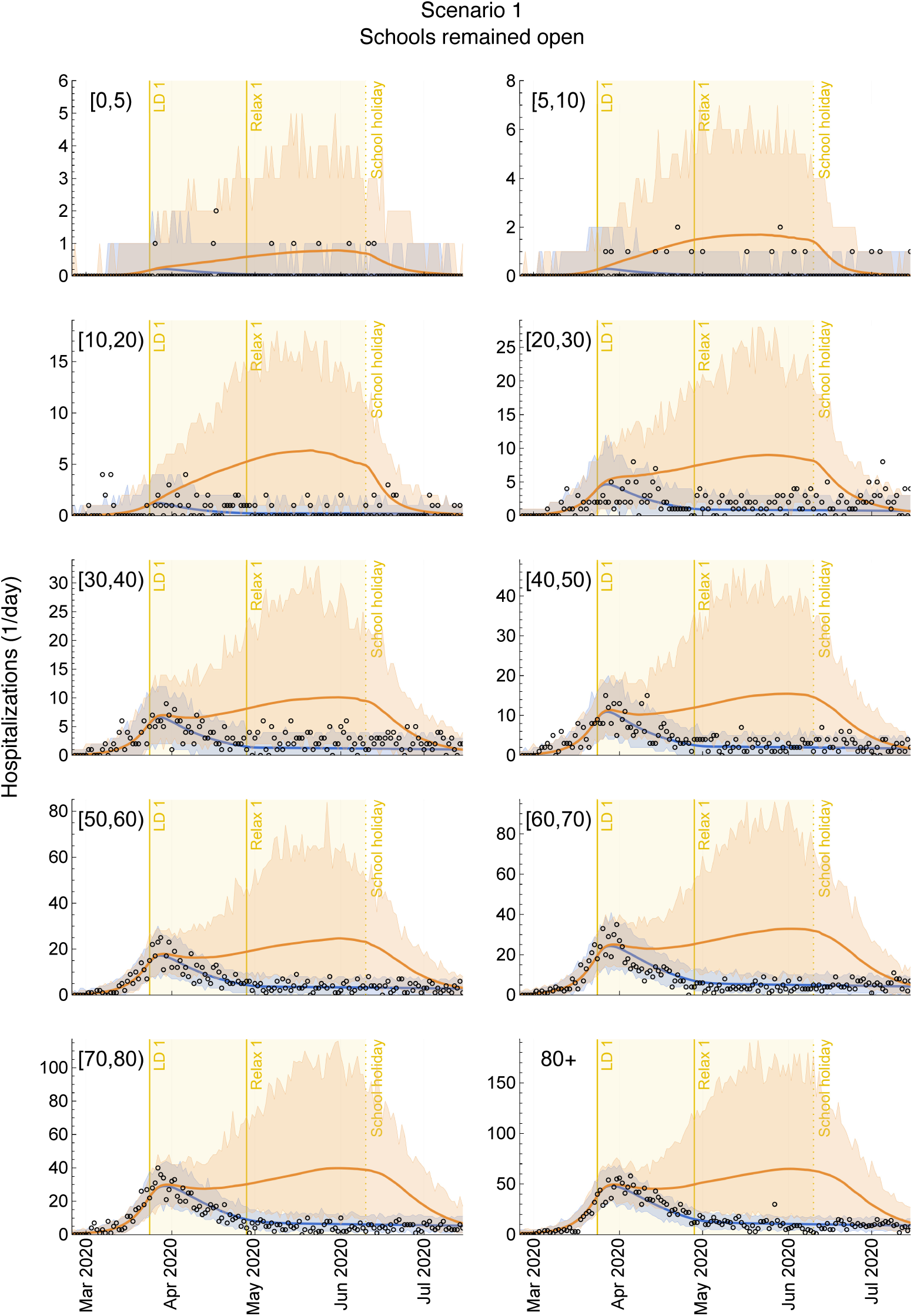
Impact of Scenario 1 on age-specific hospitalizations in ten age groups. Daily hospital admissions with COVID-19 in Scenario 1: schools remained open instead of closing during the first lockdown in spring 2020 for the ten age groups, as annotated. The circles denote daily hospital admission data. The blue lines are the median trajectories estimated from the model and the blue-shaded regions correspond to the 95% CrI of the posterior predictive distribution. The orange lines and the orange-shaded regions are the respective quantities for Scenario 1. The yellow-shaded area represents the period when schools were open in the scenario analysis. LD: lockdown. Relax: relaxation of measures.

**Figure S10.**
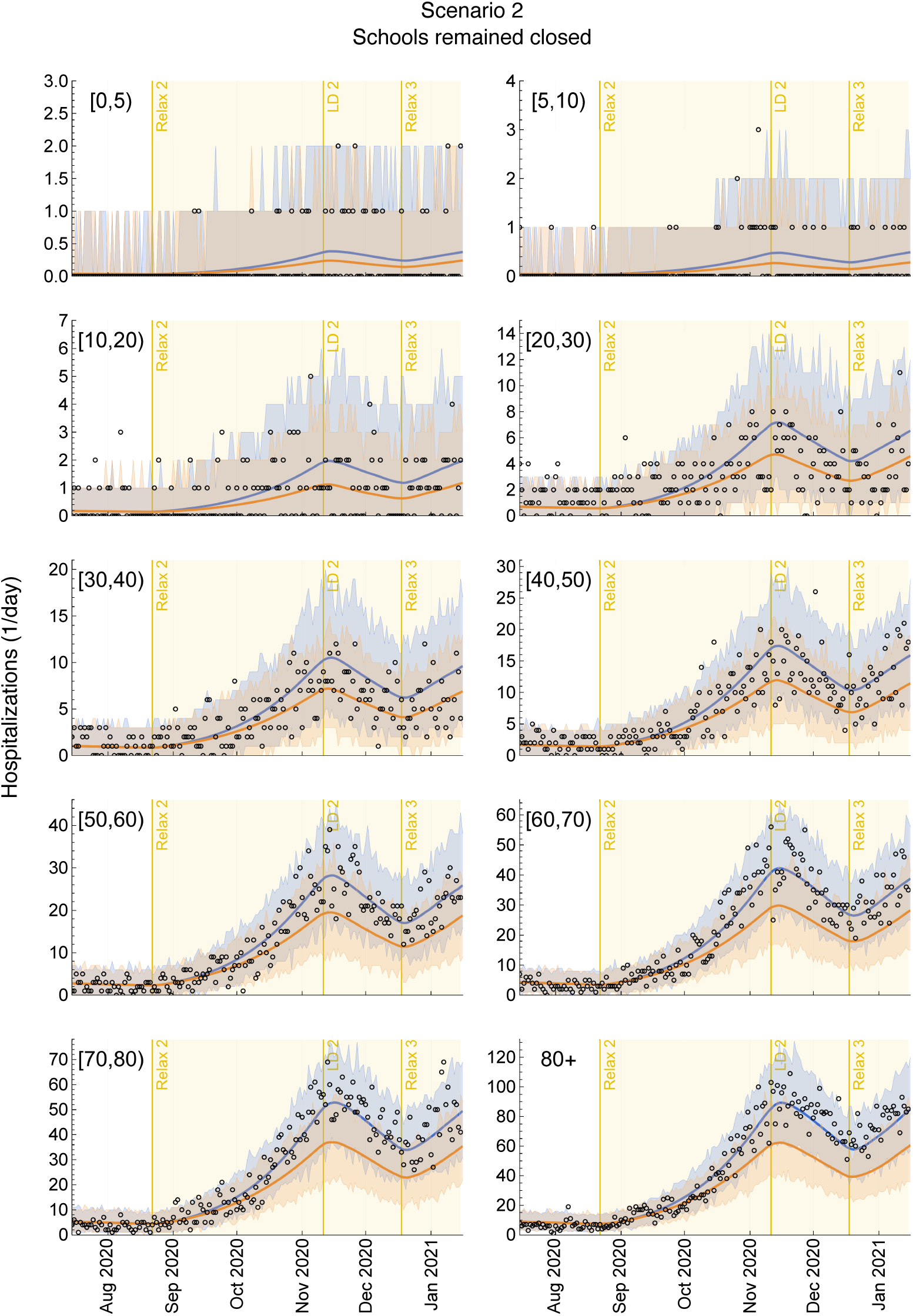
Impact of Scenario 2 on age-specific hospitalizations in ten age groups. Daily hospital admissions with COVID-19 in Scenario 2: schools remained closed instead of reopening after the 2020 summer holidays for the ten age groups, as annotated. The circles denote daily hospital admission data. The blue lines are the median trajectories estimated from the model and the blue-shaded regions correspond to the 95% CrI of the posterior predictive distribution. The orange lines and the orange-shaded regions are the respective quantities for Scenario 2. The yellow-shaded area represents the period when schools were closed in the scenario analysis. LD: lockdown. Relax: relaxation of measures.

## 7 Age-specific contributions to ***R*(*t*)**

**Figure S11.**
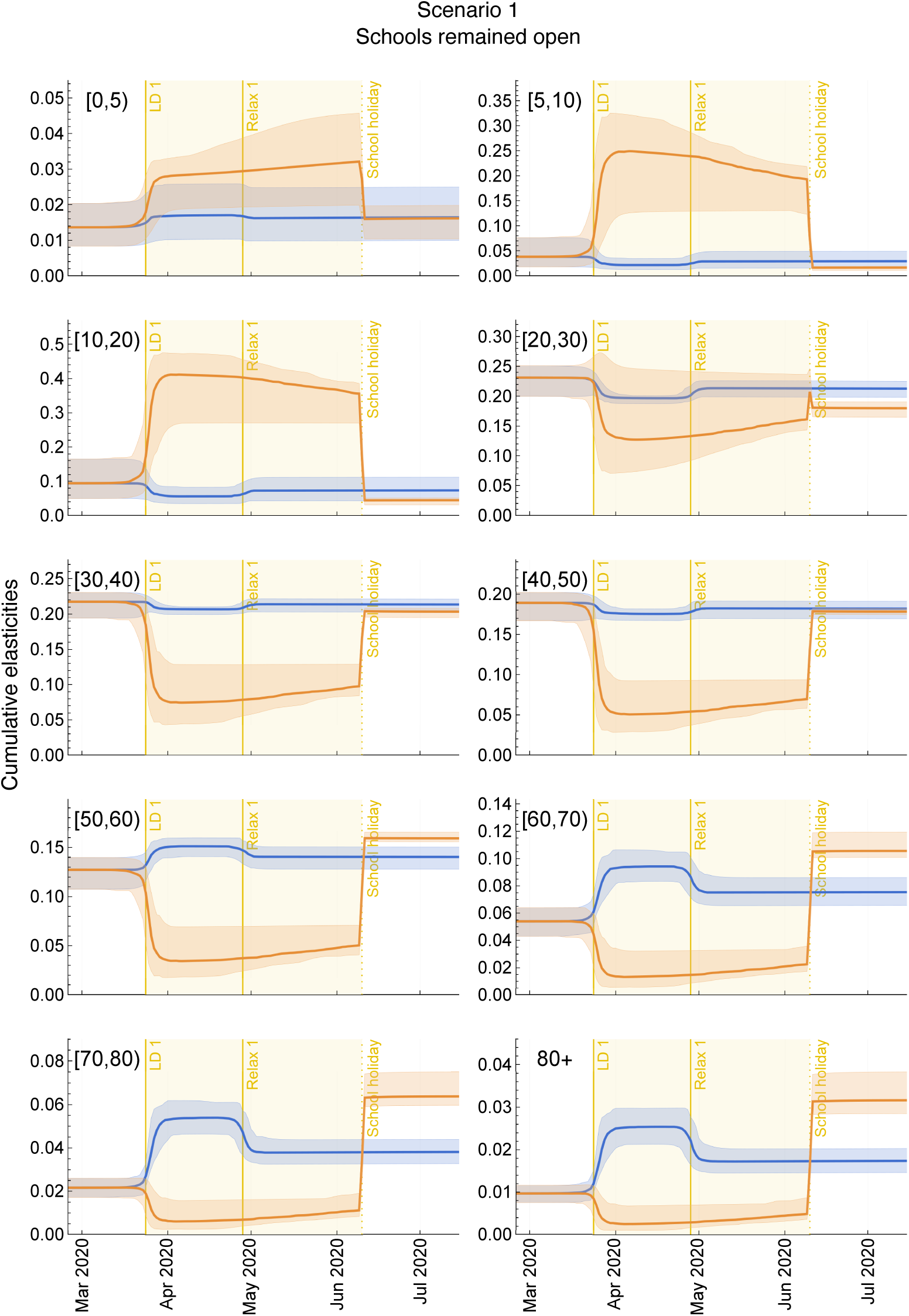
Impact of Scenario 1 on the contribution of ten age groups to. *R*(*t*). Cumulative elasticities in Scenario 1: schools remained open instead of closing during the first lockdown in spring 2020 for the ten age groups, as annotated. The blue lines are the median trajectories estimated from the model and the blue-shaded regions correspond to the 95% CrI of the posterior predictive distribution. The orange lines and the orange-shaded regions are the respective quantities for Scenario 1. The yellow-shaded area represents the period when schools were open in the scenario analysis. LD: lockdown. Relax: relaxation of measures.

**Figure S12.**
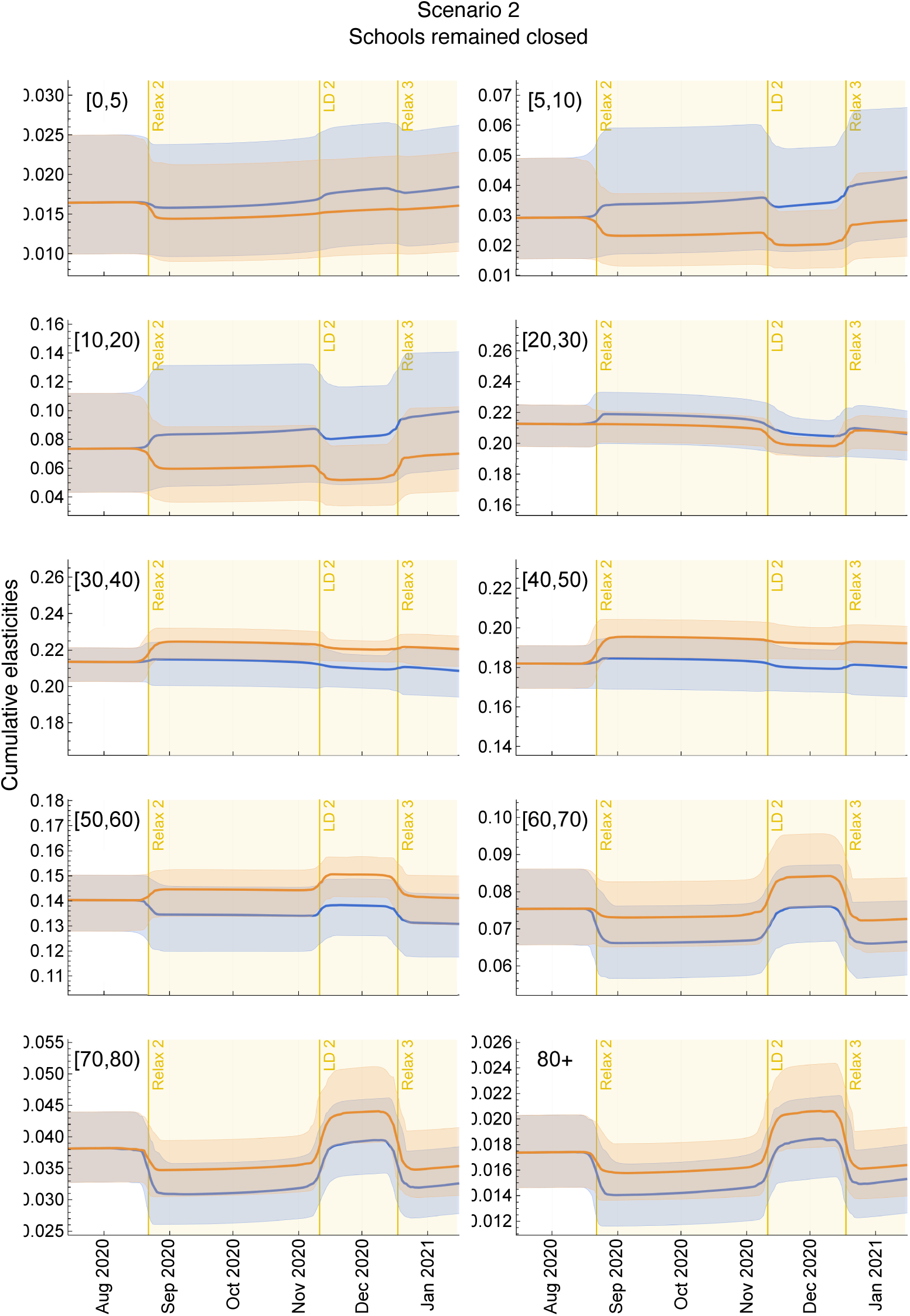
Impact of Scenario 2 on the contribution of ten age groups to *R*(*t*). Cumulative elasticities in Scenario 2: schools remained closed instead of reopening after the 2020 summer holidays for the ten age groups, as annotated. The blue lines are the median trajectories estimated from the model and the blue-shaded regions correspond to the 95% CrI of the posterior predictive distribution. The orange lines and the orange-shaded regions are the respective quantities for Scenario 2. The yellow-shaded area represents the period when schools were closed in the scenario analysis. LD: lockdown. Relax: relaxation of measures.

## 8 Comparison of cumulative hospitalizations

**Figure S13.**
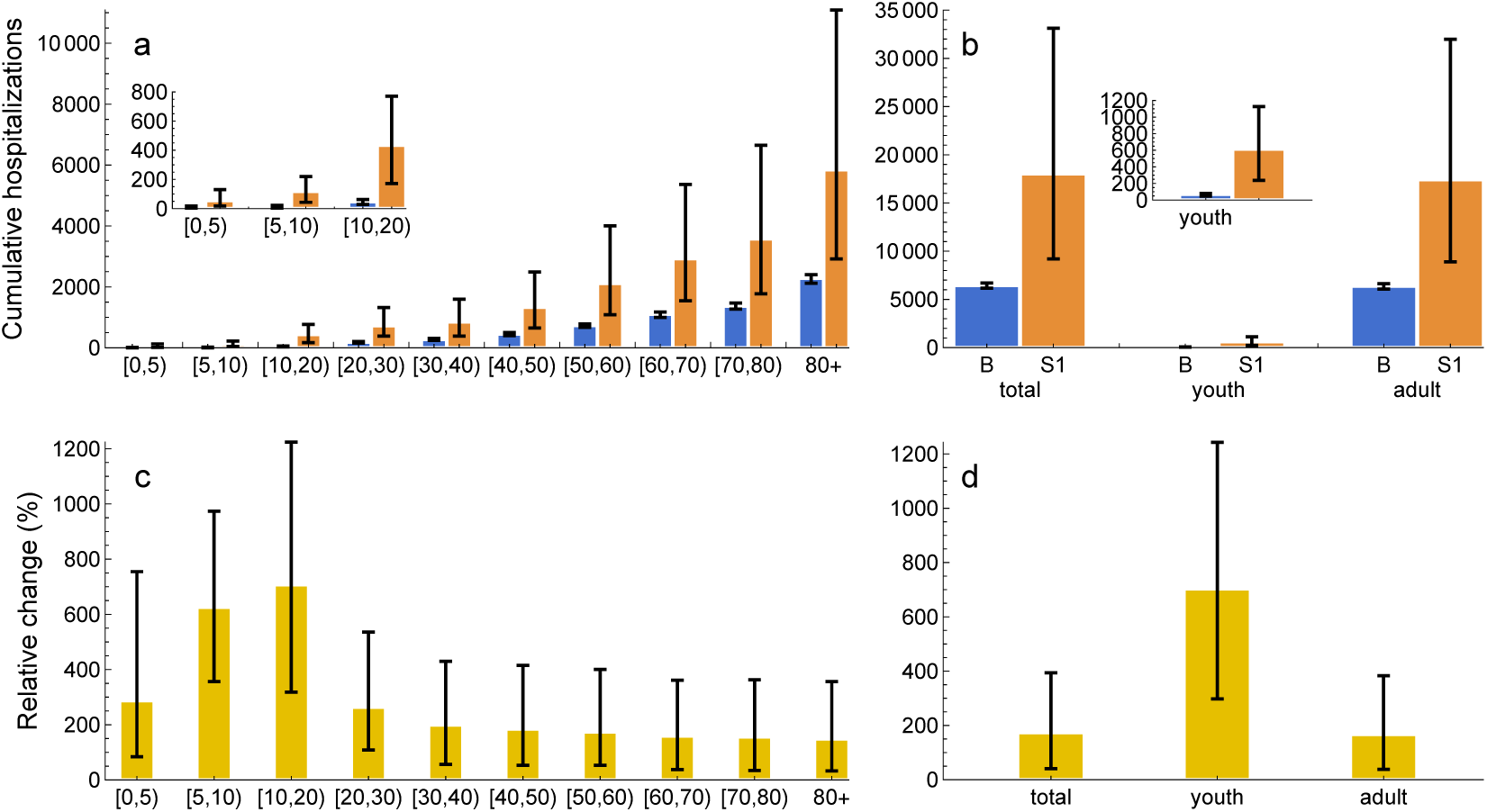
Comparison of cumulative hospitalizations for Scenario 1 vs baseline. (**a**) Age-specific, (**b**) total, youth, and adult cumulative hospital admissions with COVID-19 in Scenario 1: schools remained open instead of closing during the first lockdown in spring 2020. The bar graphs are the median values obtained on July 15, 2020 and the error bars correspond to the 95% CrI of the posterior predictive distribution. The insets for (**a**) and (**b**) show a close-up of the youth cumulative hospitalizations. The increase in cumulative hospitalizations for Scenario 1 relative to the baseline is shown for (**c**) age-specific, and (**d**) total, youth, and adult cumulative hospitalizations. B: baseline. S1: Scenario 1.

**Figure S14.**
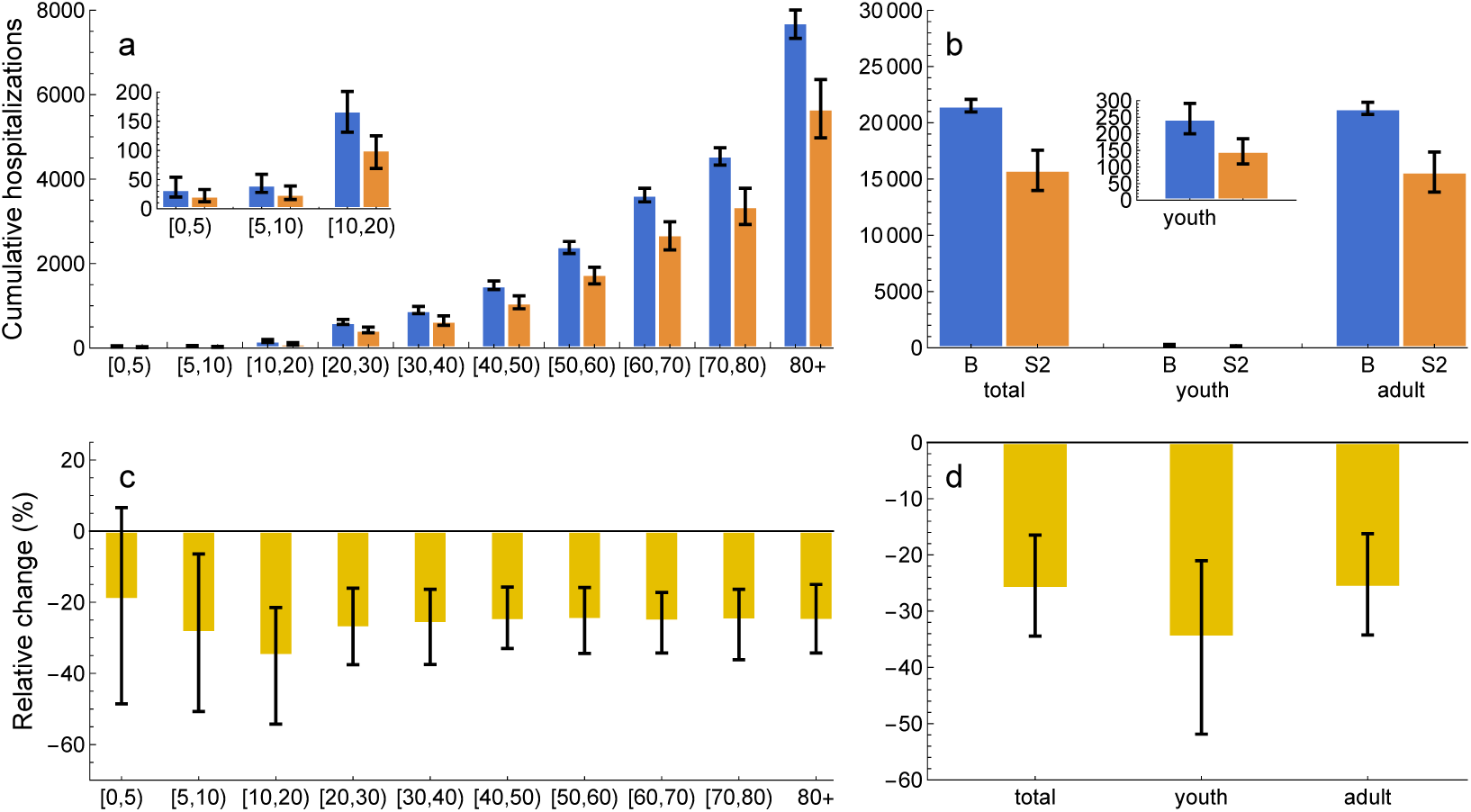
Comparison of cumulative hospitalizations for Scenario 2 vs baseline. (**a**) Age-specific, (**b**) total, youth, and adult cumulative hospital admissions with COVID-19 in Scenario 2: schools remained closed instead of reopening after the 2020 summer holidays. The bar graphs are the median values obtained on January 15, 2020 and the error bars correspond to the 95% CrI of the posterior predictive distribution. The insets for (**a**) and (**b**) show a close-up of the youth cumulative hospitalizations. The decrease in cumulative hospitalizations for Scenario 2 relative to the baseline is shown for (**c**) age-specific, and (**d**) total, youth, and adult cumulative hospitalizations. B: baseline. S2: Scenario 2.

## 9 Model projections for seroprevalence

**Figure S15.**
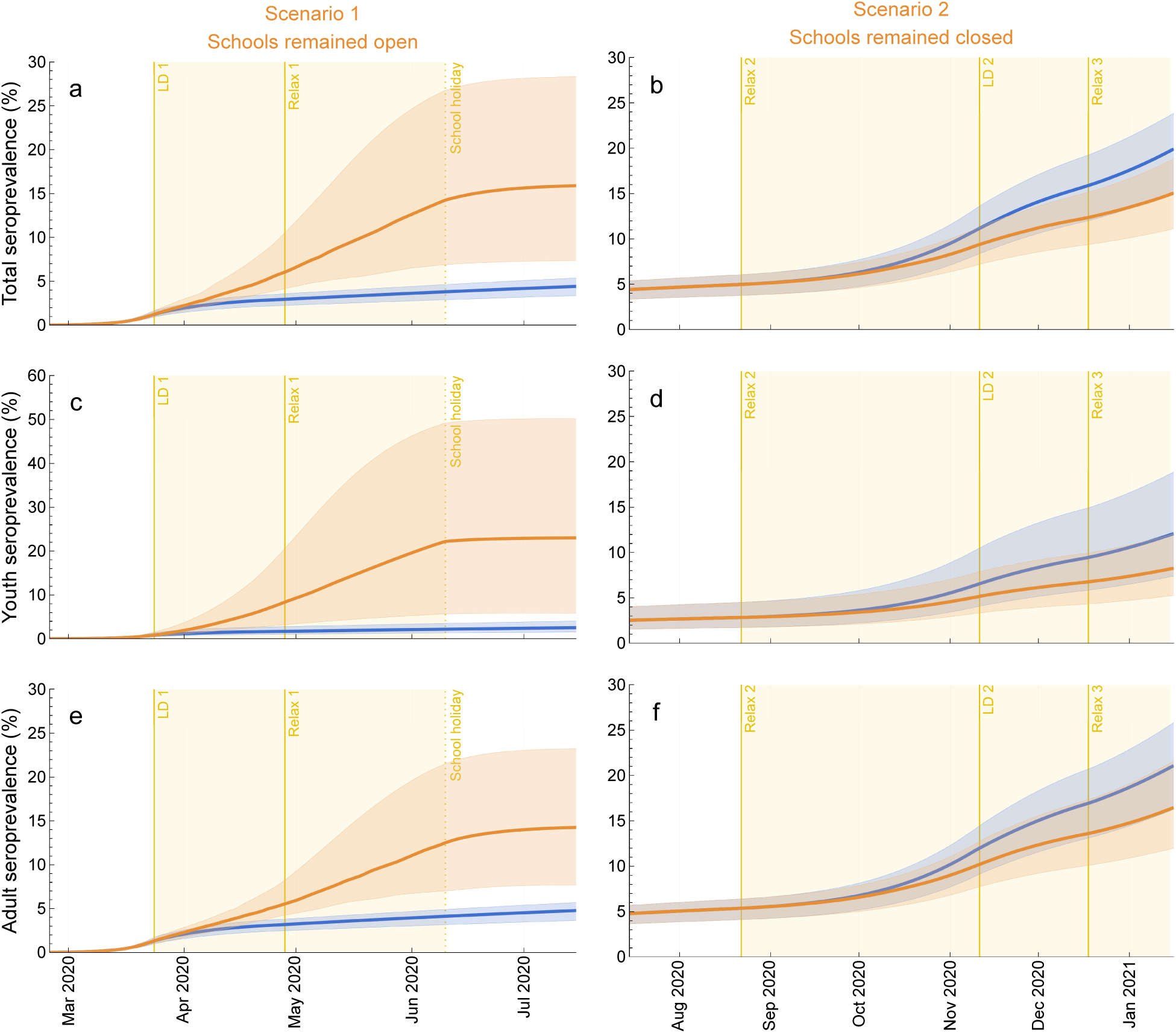
Impact of scenarios on seroprevalence. (**a**, **b**) Total, (**c**, **d**) youth (0-20 years), and (**e**, **f**) adult (20+ years) seroprevalence. (**a**, **c**, **e**) Scenario 1: schools remained open instead of closing during the first lockdown in spring 2020. (**b**, **d**, **f**) Scenario 2: schools remained closed instead of reopening after the 2020 summer holidays. The blue lines are the median trajectories estimated from the model and the blue-shaded regions correspond to the 95% CrI of the posterior predictive distribution. The orange lines and the orange-shaded regions are the respective quantities for Scenarios 1 and 2. The yellow-shaded area represents the period when schools were open (**a**, **c**, **e**) and closed (**b**, **d**, **f**) in the scenario analyses 1 and 2, respectively. LD: lockdown. Relax: relaxation of measures.

**Figure S16.**
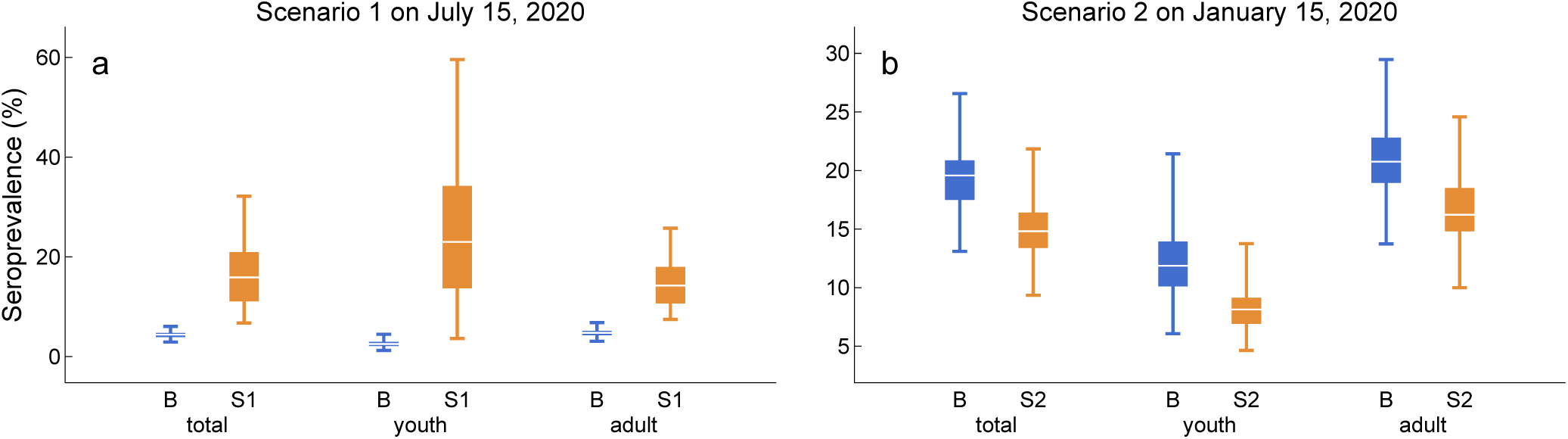
Comparison of seroprevalence on scenario enddates. Total, youth (0-20 years), and adult (20+ years) seroprevalence. (**a**) Scenario 1: schools remained open instead of closing during the first lockdown in spring 2020. (**b**) Scenario 2: schools remained closed instead of reopening after the 2020 summer holidays. The box plots represent the marginal posterior distribution of the seroprevalence on (**a**) July 15, 2020 and (**b**) January 15, 2020. B: baseline. S1: Scenario 1. S2: Scenario 2.

## 10 Uncertainty in the model projections

**Figure S17.**
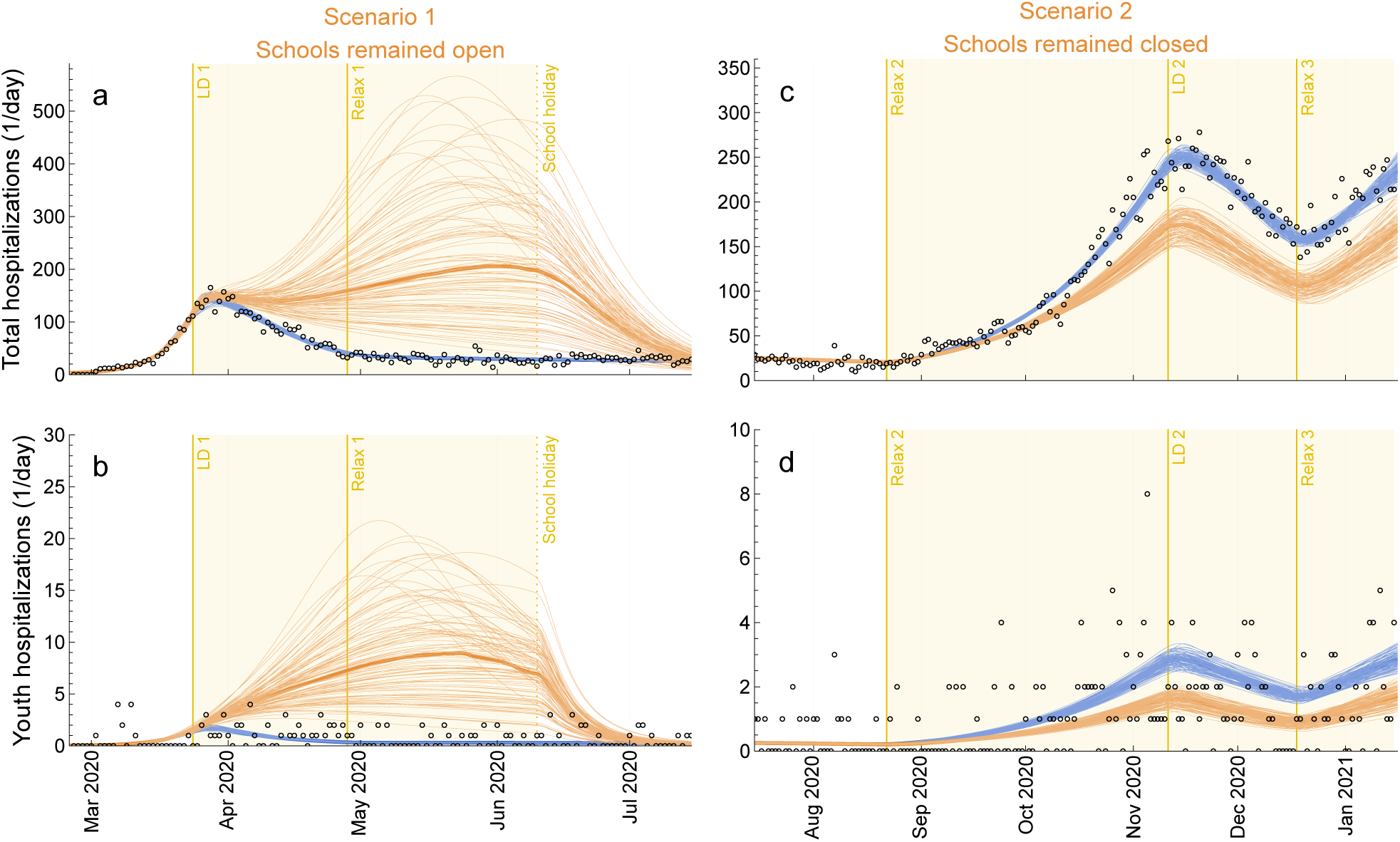
Uncertainty in the model projections. (**a**, **c**) Total and (**b**, **d**) youth (0-20 years) daily hospital admissions with COVID-19. (**a**, **b**) Scenario 1: schools remained open instead of closing during the first lockdown in spring 2020. (**c**, **d**) Scenario 2: Schools remained closed instead of reopening after the 2020 summer holidays. The circles denote daily hospital admission data. The blue lines are the 100 individual trajectories estimated from the model and the orange lines are the 100 individual trajectories of the respective quantities for Scenarios 1 and 2. Note that, for clarity, we show solutions of the ordinary differential equations without applying the observation model. The yellow-shaded area represents the period when schools were open (**a**, **b**) and closed (**c**, **d**) in the scenario analyses 1 and 2, respectively. LD: lockdown. Relax: relaxation of measures.

## 11 Sensitivity analyses for Scenario 1

### Scenario 1.1: Relaxation of measures in late May 2020

In the main analyses, Scenario 1 assumes that the strict lockdown affecting non-school contacts, represented by the contact matrix *ζa_kl_*, is maintained throughout due to the large wave of hospitalizations resulting from schools remaining open. In Scenario 1.1, described by Eq. (12), we instead assume that the strict lockdown is lifted in late April 2020, allowing the relaxation of measures affecting non-school contacts, despite the ongoing hospitalization wave in Scenario 1. The results for this scenario are shown in Supplementary Figure S18. As expected, the hospitalization wave in Scenario 1.1 is even larger than in Scenario 1.

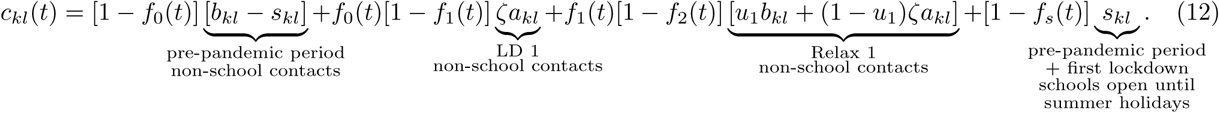

### Scenario 1.2: Additional school measures while schools stay open

In the main analyses, Scenario 1 assumes that schools remained open with pre-pandemic levels of school contacts. This assumption is justified by the abrupt onset of the pandemic in early 2020, which likely left insufficient time to implement effective mitigation measures within schools. As a result, the marginal benefit of closing schools was likely greater during this period. In Scenario 1.2, described by Eq. (13), we relaxed this assumption by applying a mitigation scaling factor *ζ*_1_ = *ζ* to the pre-pandemic school contact matrix, *s_kl_*. This assumption reflects a scenario in which, during the first lockdown, the reduction in school contacts due to within-school mitigation measures among teachers and students was the same as the reduction for the non-school contacts due to protective measures (i.e., mask-wearing, refraining from shaking hands, physical distancing, etc.) in the general population. The results for this alternative scenario are shown in Supplementary Figure S19. We find that the resulting hospitalization wave exceeds the observed wave in the data. If school-based contacts are reduced by a large amount, the deviation would be smaller. This is expected, as the limiting case of *ζ*_1_ = 0 corresponds to complete school closure.

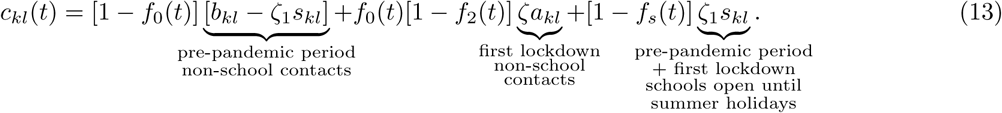

**Figure S18.**
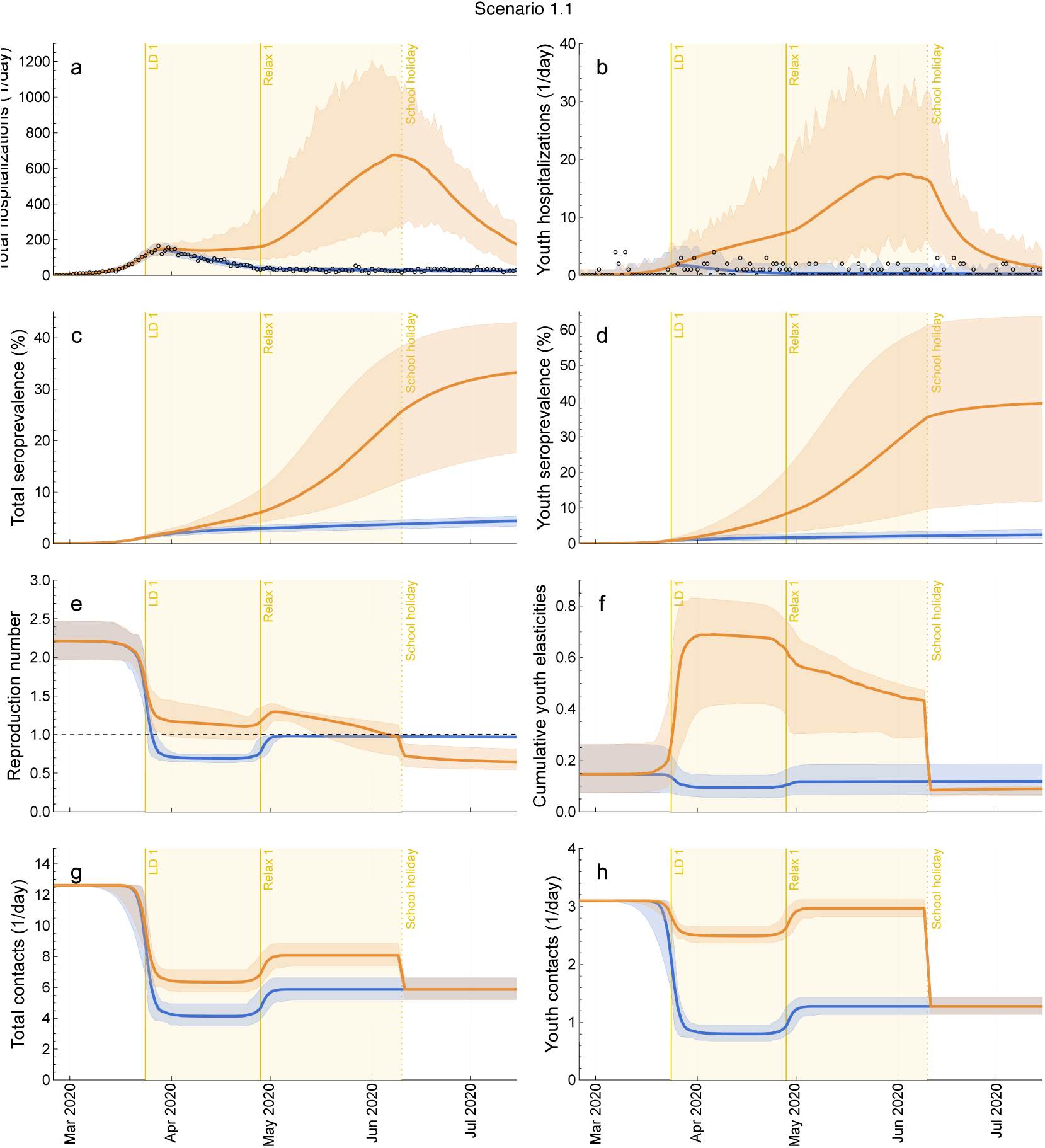
Impact of Scenario 1.1 on model outcome measures. (**a**) Total and (**b**) youth hospitalizations, (**c**) total and (**d**) youth seroprevalence, (**e**) time-varying reproduction number, (**f**) relative contribution of youth to *R*(*t*), and (**g**) total and (**h**) youth contacts for Scenario 1.1: relaxation of measures in late May 2020. The circles denote daily hospital admission data. The dashed line corresponds to *R*(*t*) = 1. The blue lines are the median trajectories estimated from the model and the blue-shaded regions correspond to the 95% CrI of the posterior predictive distribution. The orange lines and the orange-shaded regions are the respective quantities for Scenario 1.1. The yellow-shaded area represents the period when schools were open in the scenario analysis. LD: lockdown. Relax: relaxation of measures.

**Figure S19.**
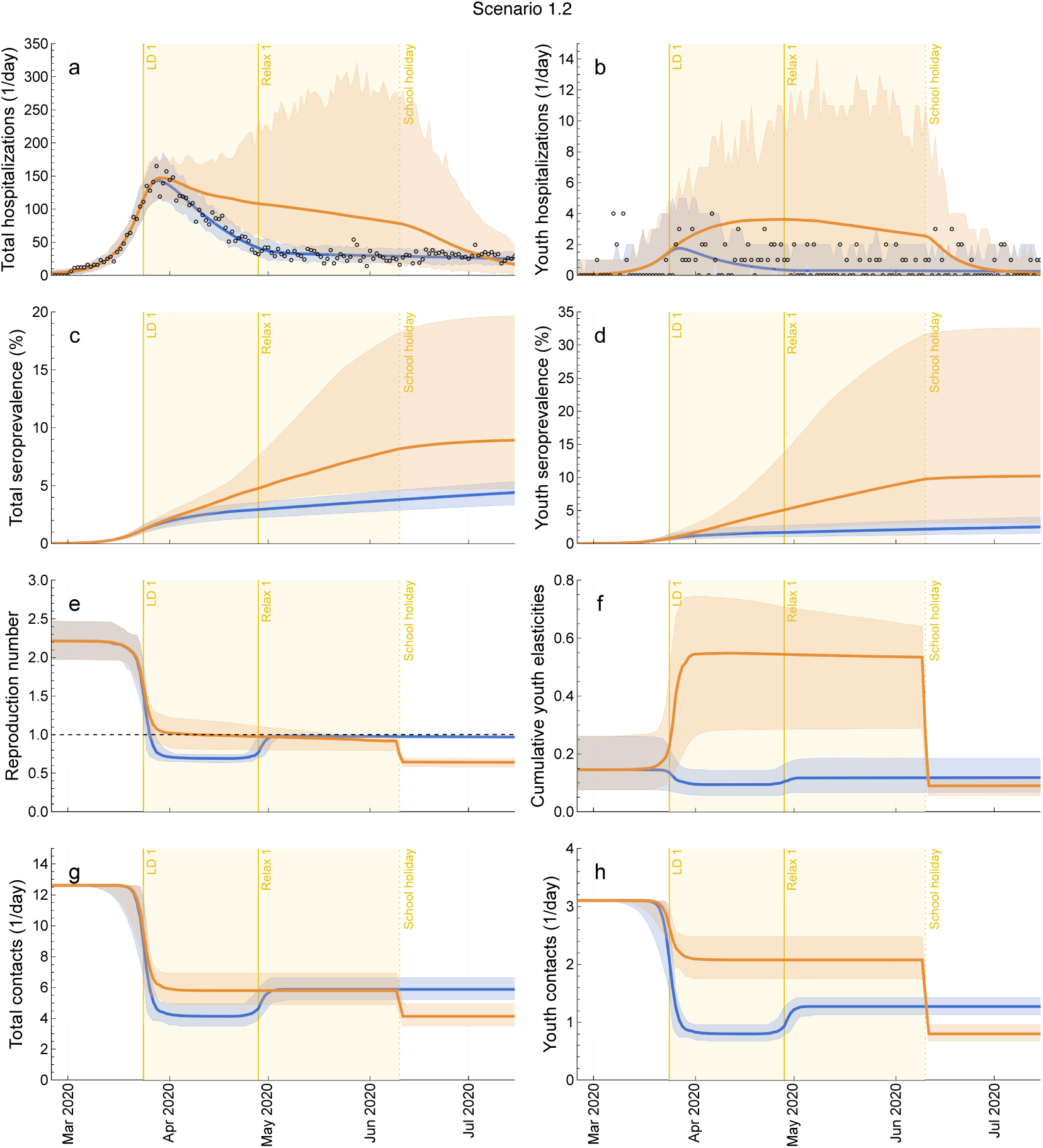
Impact of Scenario 1.2 on model outcome measures. (**a**) Total and (**b**) youth hospitalizations, (**c**) total and (**d**) youth seroprevalence, (**e**) time-varying reproduction number, (**f**) relative contribution of youth to *R*(*t*), and (**g**) total and (**h**) youth contacts for Scenario 1.2: additional school measures while schools stay open. The circles denote daily hospital admission data. The dashed line corresponds to *R*(*t*) = 1. The blue lines are the median trajectories estimated from the model and the blue-shaded regions correspond to the 95% CrI of the posterior predictive distribution. The orange lines and the orange-shaded regions are the quantities for Scenario 1.2. The yellow-shaded area represents the period when schools were open in the scenario analysis. LD: lockdown. Relax: relaxation of measures.

## 12 Sensitivity analyses for Scenario 2

### Scenario 2.1: School winter holidays

In the main analyses, Scenario 2 does not account for school closures due to the winter holidays. In Scenario 2.1, defined by Eq. (14), we assume that schools are closed during the final period, Relax 3, which begins in mid-December 2020 and coincides with the start of the winter holidays. The results are shown in Supplementary Figure S20. The impact of school holidays on hospitalizations is small.

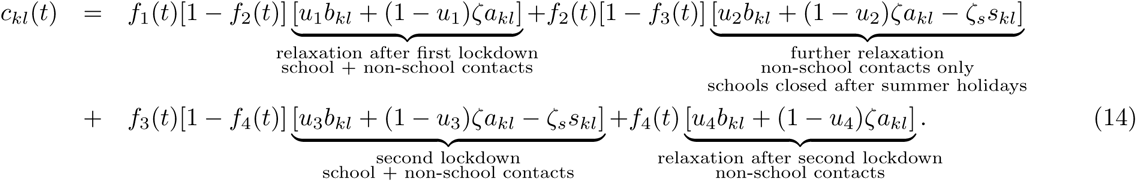

### Scenario 2.2: Alternative reductions in school-based contacts

In the main analyses, the calibrated parameter *ζ_s_*, representing the mitigation factor in school-based contacts, is applied to school contact matrix across all three periods in Scenario 2. This reflects the fact that no major changes occurred in within-school measures during the entire Scenario 2 period. In the alternative Scenario 2.2, defined by Eq. (15), we instead use the parameters *u_j_* for each period to represent the fraction of school contacts removed due to school closures. The results are shown in Supplementary Figure S21. Compared to Scenario 2 in the main text, this scenario results in a somewhat smaller epidemic wave. However, the overall qualitative behavior of the model remains similar. School closures can mitigate, but not prevent, the autumn 2020 wave.

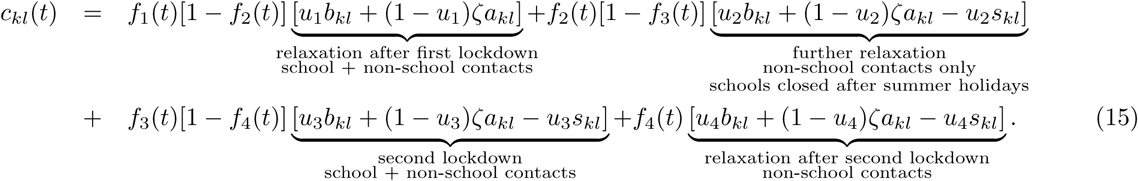

### Scenario 2.3: Continued Scenario 1

We continue Scenario 1 into the period of Scenario 2 by implementing the same measures as that of the baseline during the second wave. For the contact formulation, Scenario 2.3 basically considered Equation 3 and Equation 4. The results are shown in Figure S22.

**Figure S20.**
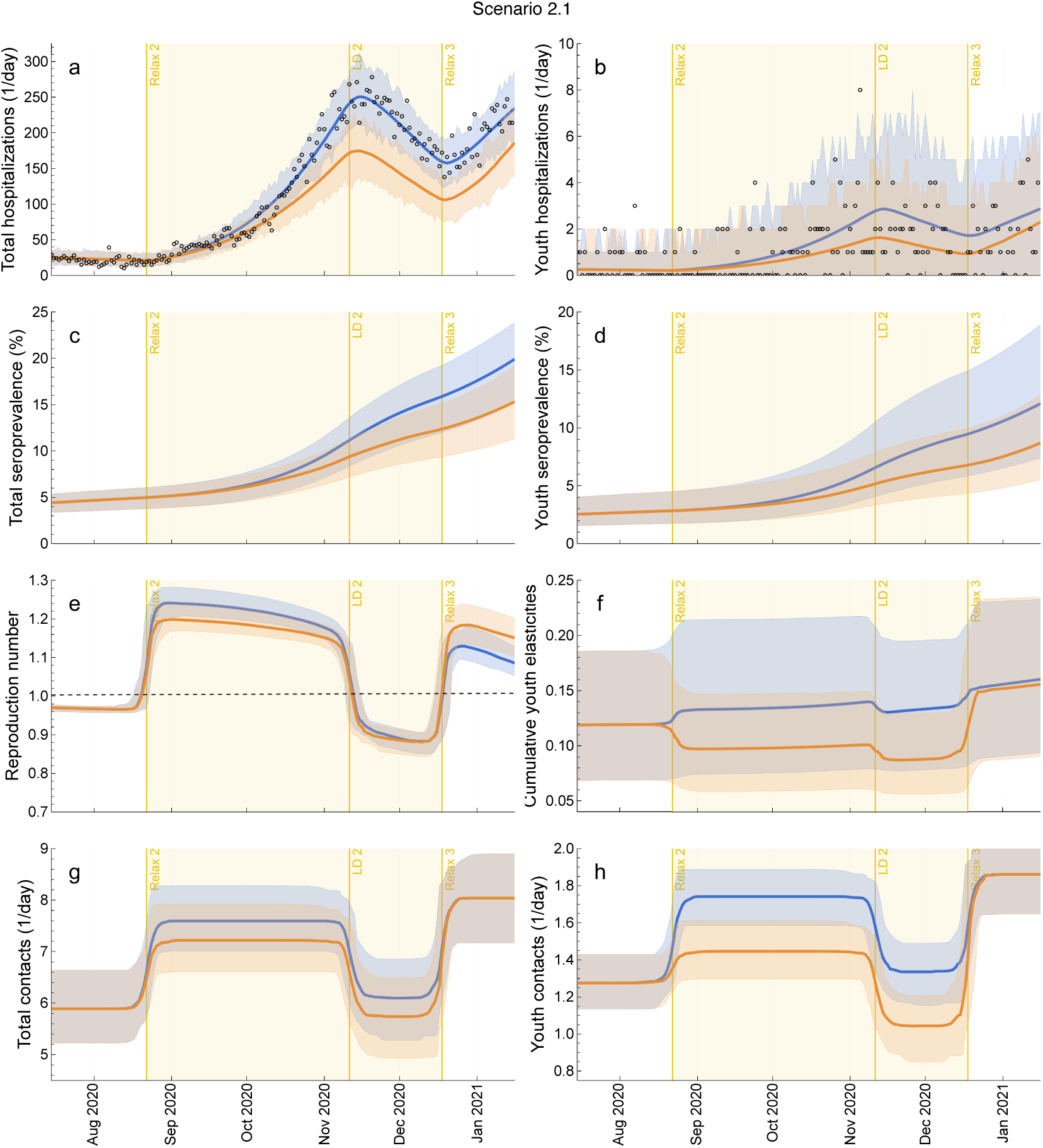
Impact of Scenario 2.1 on model outcome measures. (**a**) Total and (**b**) youth hospitalizations, (**c**) total and (**d**) youth seroprevalence, (**e**) time-varying reproduction number, (**f**) relative contribution of youth to *R*(*t*), and (**g**) total and (**h**) youth contacts for Scenario 2.1: school winter holidays. The circles denote daily hospital admission data. The dashed line corresponds to *R*(*t*) = 1. The blue lines are the median trajectories estimated from the model and the blue-shaded regions correspond to the 95% CrI of the posterior predictive distribution. The orange lines and the orange-shaded regions are the respective quantities for Scenario 2.1. The yellow-shaded area represents the period when schools were closed in the scenario analysis. LD: lockdown. Relax: relaxation of measures.

**Figure S21.**
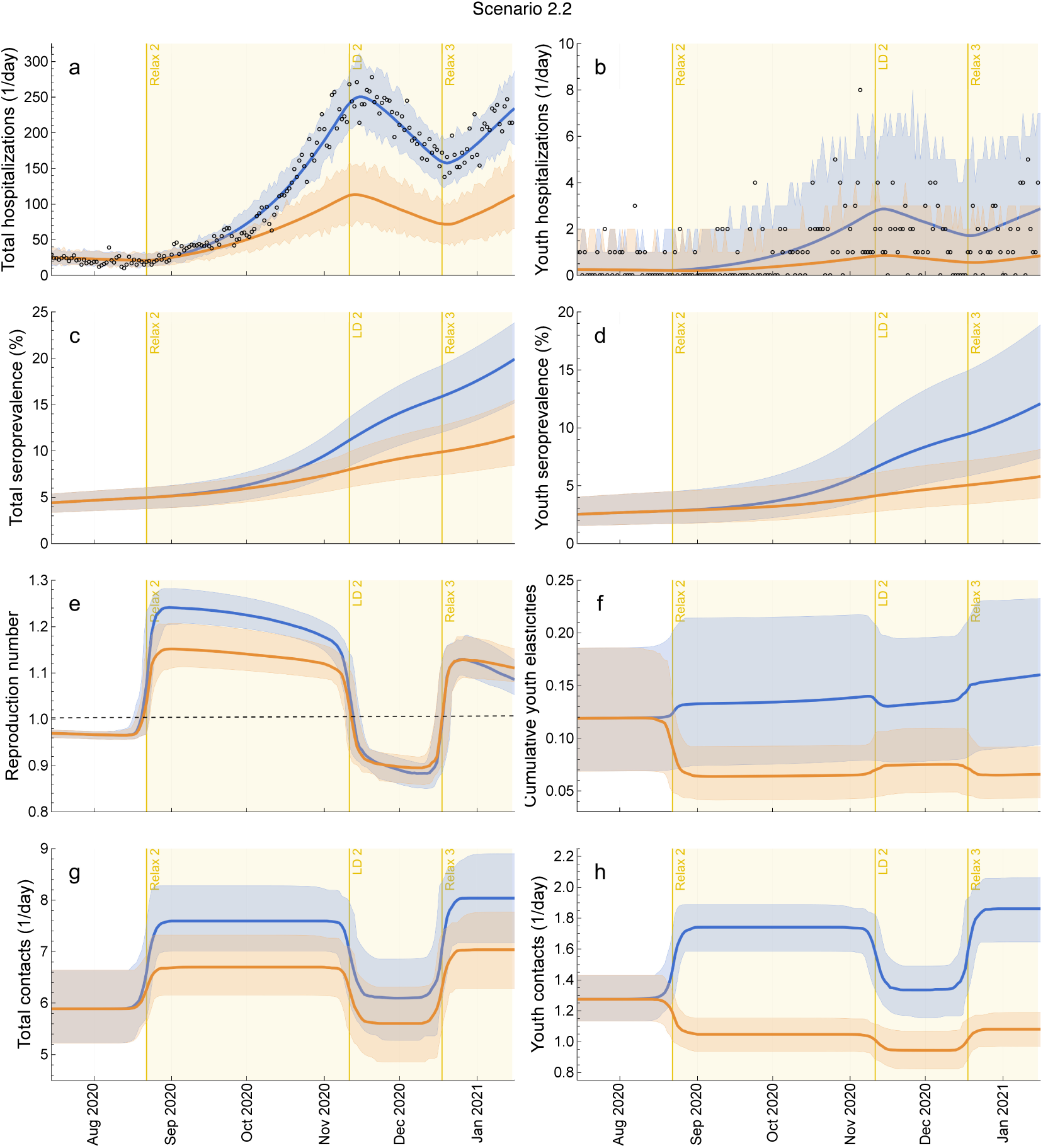
Impact of Scenario 2.2 on model outcome measures. (**a**) Total and (**b**) youth hospitalizations, (**c**) total and (**d**) youth seroprevalence, (**e**) time-varying reproduction number, (**f**) relative contribution of youth to *R*(*t*), and (**g**) total and (**h**) youth contacts for Scenario 2.2: alternative reductions in school-based contacts. The circles denote daily hospital admission data. The dashed line corresponds to *R*(*t*) = 1. The blue lines are the median trajectories estimated from the model and the blue-shaded regions correspond to the 95% CrI of the posterior predictive distribution. The orange lines and the orange-shaded regions are the respective quantities for Scenario 2.2. The yellow-shaded area represents the period when schools were closed in the scenario analysis. LD: lockdown. Relax: relaxation of measures.

**Figure S22.**
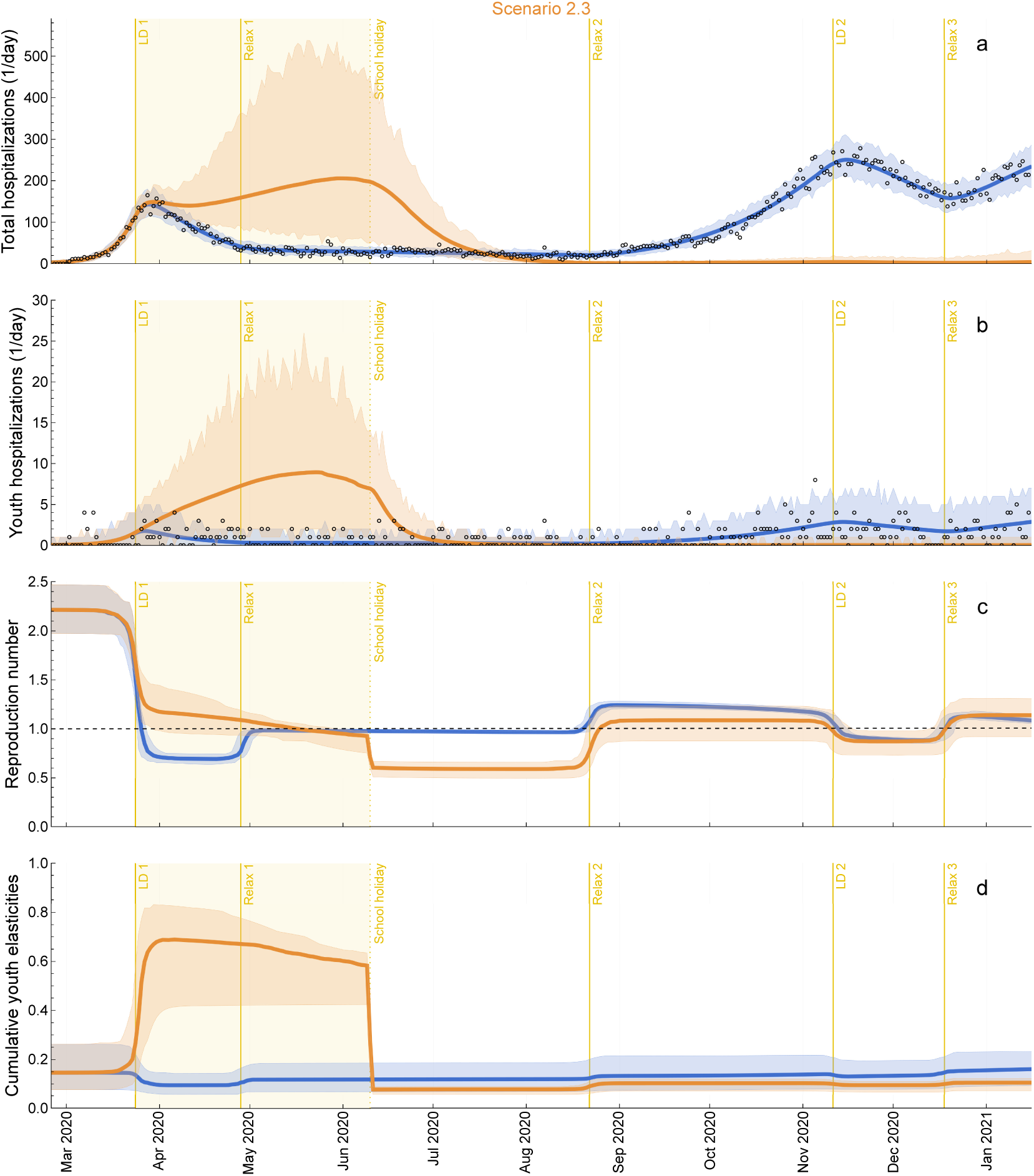
Impact of Scenario 2.3 on model outcome measures. (**a**) Total and (**b**) youth hospitalizations, (**c**) time-varying reproduction number, and (**d**) relative contribution of youth to *R*(*t*) for Scenario 2.3: continued Scenario 1. The circles denote daily hospital admission data. The dashed line corresponds to *R*(*t*) = 1. The blue lines are the median trajectories estimated from the model and the blue-shaded regions correspond to the 95% CrI of the posterior predictive distribution. The orange lines and the orange-shaded regions are the quantities for Scenario 2.3. The yellow-shaded area represents the period when schools were open in the scenario analysis. LD: lockdown. Relax: relaxation of measures.

## References

[1] Cordovil R, Ribeiro L, Moreira M, Pombo A, Rodrigues LP, Luz C, et al. Effects of the COVID-19 pandemic on preschool children and preschools in Portugal. Journal of Physical Education and Sport. 2021;21(Supplement issue 1):492–499. doi:10.7752/jpes.2021.s1052.

[2] Lemos-Paiao AP, Silva CJ, Torres DF. A new compartmental epidemiological model for COVID-19 with a case study of Portugal. Ecological Complexity. 2020;44:100885. doi:10.1016/j.ecocom.2020.100885.

[3] Pombo A, Luz C, Rodrigues LP, Ferreira C, Cordovil R. Correlates of children’s physical activity during the COVID-19 confinement in Portugal. Public Health. 2020;189:14–19. doi:10.1016/j.puhe.2020.09.009.

[4] Torres AR, Rodrigues AP, Sousa-Uva M, Kislaya I, Silva S, Antunes L, et al. Impact of stringent non-pharmaceutical interventions applied during the second and third COVID-19 epidemic waves in Portugal, 9 November 2020 to 10 February 2021: an ecological study. Eurosurveillance. 2022;27(23):2100497. doi:10.2807/1560-7917.ES.2022.27.23.2100497.

[5] Caetano C, Morgado ML, Patrício P, Leite A, Machado A, Torres A, et al. Measuring the impact of COVID-19 vaccination and immunity waning: a modelling study for Portugal. Vaccine. 2022;40(49):7115–7121. doi:10.1016/j.vaccine.2022.10.007.

[6] Caetano C, Morgado ML, Patrício P, Pereira JF, Nunes B. Mathematical modelling of the impact of non-pharmacological strategies to control the COVID-19 epidemic in Portugal. Mathematics. 2021;9(10):1084. doi:10.3390/math9101084.

[7] Rice K, Wynne B, Martin V, Ackland GJ. Effect of school closures on mortality from coronavirus disease 2019: old and new predictions. BMJ. 2020;371. doi:10.7488/ds/2912.

[8] Rozhnova G, van Dorp CH, Bruijning-Verhagen P, Bootsma MC, van de Wijgert JH, Bonten MJ, et al. Model-based evaluation of school-and non-school-related measures to control the COVID-19 pandemic. Nature Communications. 2021;12(1):1614. doi:10.1038/s41467-021-21899-6.

[9] Walsh S, Chowdhury A, Braithwaite V, Russell S, Birch JM, Ward JL, et al. Do school closures and school reopenings affect community transmission of COVID-19? A systematic review of observational studies. BMJ Open. 2021;11(8):e053371. doi:10.1136/bmjopen-2021-053371.

[10] Littlecott H, Krishnaratne S, Burns J, Rehfuess E, Sell K, Klinger C, et al. Measures implemented in the school setting to contain the COVID-19 pandemic. The Cochrane Database of Systematic Reviews. 2024;5:CD015029–CD015029. doi:10.1002/14651858.CD015029.

[11] Panovska-Griffiths J, Kerr CC, Stuart RM, Mistry D, Klein DJ, Viner RM, et al. Determining the optimal strategy for reopening schools, the impact of test and trace interventions, and the risk of occurrence of a second COVID-19 epidemic wave in the UK: a modelling study. The Lancet Child & Adolescent Health. 2020;4(11):817–827. doi:10.1016/S2352-4642(20)30250-9.

[12] Baxter A, Oruc BE, Asplund J, Keskinocak P, Serban N. Evaluating scenarios for school reopening under COVID19. BMC Public Health. 2022;22(1):496. doi:10.1186/s12889-022-12910-w.

[13] Ragonnet R, Hughes AE, Shipman DS, Meehan MT, Henderson AS, Briffoteaux G, et al. Estimating the impact of school closures on the COVID-19 dynamics in 74 countries: A modelling analysis. PLOS Medicine. 2025;22(1):e1004512. doi:10.1371/journal.pmed.1004512.

[14] Prem K, Liu Y, Russell TW, Kucharski AJ, Eggo RM, Davies N, et al. The effect of control strategies to reduce social mixing on outcomes of the COVID-19 epidemic in Wuhan, China: a modelling study. The Lancet Public Health. 2020;5(5):e261–e270. doi:10.1016/S2468-2667(20)30073-6.

[15] Alves A, da Costa NM, Morgado P, da Costa EM. Uncovering COVID-19 infection determinants in Portugal: towards an evidence-based spatial susceptibility index to support epidemiological containment policies. International Journal of Health Geographics. 2023;22(1):8. doi:10.1186/s12942-023-00329-4.

[16] Caetano C, Angeli L, Varela-Lasheras I, Coletti P, Morgado L, Lima P, et al. Identifying the main drivers of transmission in the early phase of the COVID-19 pandemic in Portugal. Scientific Reports. 2024;14(1):30689. doi:10.1038/s41598-024-76604-6.

[17] Gomes JJF, Ferreira A, Alves A, Sequeira BN. A risk scoring model of COVID-19 at hospital admission. PloS One. 2023;18(7):e0288460. doi:10.1371/journal.pone.0288460.

[18] Milhinhos A, Costa PM. On the progression of COVID-19 in Portugal: A comparative analysis of active cases using non-linear regression. Frontiers in Public Health. 2020;8:495. doi:10.3389/fpubh.2020.00495.

[19] Pinto CM, Tenreiro Machado J, Burgos-Simón C. Modified SIQR model for the COVID-19 outbreak in several countries. Mathematical Methods in the Applied Sciences. 2024;47(5):3273–3288. doi:10.1002/mma.8082.

[20] Tedim S, Afreixo V, Felgueiras M, Leitao RP, Pinheiro SJ, Silva CJ. Evaluating COVID-19 in Portugal: Bootstrap confidence interval. AIMS Mathematics. 2024;9(2):2756 – 2765. doi:10.3934/math.2024136.

[21] Pais RJ, Taveira N. Predicting the evolution and control of the COVID-19 pandemic in Portugal. F1000Research. 2020;9. doi:10.12688/f1000research.23401.2.

[22] Machado B, Antunes L, Caetano C, Pereira JF, Nunes B, Patrício P, et al. The impact of vaccination on the evolution of COVID-19 in Portugal. Mathematical Biosciences and Engineering. 2022;19:936–952. doi:10.3934/mbe.2022043.

[23] Viana J, van Dorp CH, Nunes A, Gomes MC, van Boven M, Kretzschmar ME, et al. Controlling the pandemic during the SARS-CoV-2 vaccination rollout. Nature Communications. 2021;12(1):3674. doi:10.1038/s41467-021-23938-8.

[24] Esquível ML, Krasii NP, Guerreiro GR, Patrício P. The multi-compartment SI (RD) model with regime switching: An application to COVID-19 pandemic. Symmetry. 2021;13(12):2427. doi:10.3390/sym13122427.

[25] Silva CJ, Cruz C, Torres DF, Munuzuri AP, Carballosa A, Area I, et al. Optimal control of the COVID-19 pandemic: controlled sanitary deconfinement in Portugal. Scientific Reports. 2021;11(1):3451. doi:10.1038/s41598-021-83075-6.

[26] Balsa C, Lopes I, Guarda T, Rufino J. Computational simulation of the COVID-19 epidemic with the SEIR stochastic model. Computational and Mathematical Organization Theory. 2023; p. 1–19. doi:10.1007/s10588-021-09327-y.

[27] Figueira JR, Oliveira HM, Serro AP, Colaço R, Froes F, Cordeiro CR, et al. A multiple criteria approach for building a pandemic impact assessment composite indicator: The case of COVID-19 in Portugal. European Journal of Operational Research. 2023;309(2):795–818. doi:10.1016/j.ejor.2023.01.025.

[28] Rosa S, Torres DF. Fractional modelling and optimal control of COVID-19 transmission in Portugal. Axioms. 2022;11(4):170. doi:10.3390/axioms11040170.

[29] Beira MJ, Sebastião PJ. A differential equations model-fitting analysis of COVID-19 epidemiological data to explain multi-wave dynamics. Scientific Reports. 2021;11(1):16312. doi:10.1038/s41598-021-95494-6.

[30] Kislaya I, Gonçalves P, Barreto M, Sousa Rd, Garcia AC, Matos R, et al. Seroprevalence of SARS-CoV-2 infection in Portugal in May-July 2020: results of the first national serological survey (ISNCOVID-19). Acta Medica Portuguesa. 2021;34(2):87–94. doi:10.20344/amp.15122.

[31] PORDATA. Base de Dados Portugal Contemporâneo; 2024. Available from: https://www.pordata.pt/pt.

[32] Mistry D, Litvinova M, Pastore y Piontti A, Chinazzi M, Fumanelli L, Gomes MF, et al. Inferring high-resolution human mixing patterns for disease modeling. Nature Communications. 2021;12(1):323. doi:10.1038/s41467-020-20544-y.

[33] Backer JA, Mollema L, Vos ER, Klinkenberg D, Van Der Klis FR, De Melker HE, et al. Impact of physical distancing measures against COVID-19 on contacts and mixing patterns: repeated cross-sectional surveys, the Netherlands, 2016–17, April 2020 and June 2020. Eurosurveillance. 2021;26(8):2000994. doi:10.2807/1560-7917.ES.2021.26.8.2000994.

[34] Gabinetes do Secretário de Estado Adjunto e da Educação e da Secretária de Estado da Educação. Díario da República, 2.ª série — N.° 128 — 3 de julho de 2020; 2020. Available from: https://files.diariodarepublica.pt/2s/2020/07/128000002/0000400009.pdf.

[35] Carpenter B, Gelman A, Hoffman M, Lee D, Goodrich B, Betancourt M, et al. Stan: A probabilistic programming language. Journal of Statistical Software. 2017;76(1):1–32. 10.18637/jss.v076.i01.

[36] Lauer SA, Grantz KH, Bi Q, Jones FK, Zheng Q, Meredith HR, et al. The incubation period of coronavirus disease 2019 (COVID-19) from publicly reported confirmed cases: estimation and application. Annals of Internal Medicine. 2020;172(9):577–582. doi:10.7326/M20-050.

[37] Park M, Cook AR, Lim JT, Sun Y, Dickens BL. A systematic review of COVID-19 epidemiology based on current evidence. Journal of Clinical Medicine. 2020;9(4):967. doi:10.3390/jcm9040967.

[38] Ali ST, Wang L, Lau EH, Xu XK, Du Z, Wu Y, et al. Serial interval of SARS-CoV-2 was shortened over time by nonpharmaceutical interventions. Science. 2020;369(6507):1106–1109. doi:10.1126/science.abc9004.

[39] Jing QL, Liu MJ, Zhang ZB, Fang LQ, Yuan J, Zhang AR, et al. Household secondary attack rate of COVID-19 and associated determinants in Guangzhou, China: a retrospective cohort study. The Lancet Infectious Diseases. 2020;20(10):1141–1150. 10.1016/S1473-3099(20)30471-0.

[40] Diekmann O, Heesterbeek J, Roberts MG. The construction of next-generation matrices for compartmental epidemic models. Journal of the Royal Society Interface. 2010;7(47):873–885. doi:10.1098/rsif.2009.0386.

[41] Caswell H. Matrix population models. vol. 1. Sinauer Sunderland, MA; 2000.

[42] Van den Driessche P. Reproduction numbers of infectious disease models. Infectious Disease Modelling. 2017;2(3):288–303. doi:10.1016/j.idm.2017.06.002.

[43] de Kroon H, Plaisier A, van Groenendael J, Caswell H. Elasticity: the relative contribution of demographic parameters to population growth rate. Ecology. 1986;67(5):1427–1431. doi:10.2307/1938700.

[44] Van Groenendael J, De Kroon H, Kalisz S, Tuljapurkar S. Loop analysis: evaluating life history pathways in population projection matrices. Ecology. 1994;75(8):2410–2415. doi:10.2307/1940894.

[45] Davies NG, Kucharski AJ, Eggo RM, Gimma A, Edmunds WJ, Jombart T, et al. Effects of non-pharmaceutical interventions on COVID-19 cases, deaths, and demand for hospital services in the UK: a modelling study. The Lancet Public Health. 2020;5(7):e375–e385. doi:10.1016/S2468-2667(20)30133-X.

[46] Mazrekaj D, De Witte K. The impact of school closures on learning and mental health of children: Lessons from the COVID-19 pandemic. Perspectives on Psychological Science. 2024;19(4):686–693. doi:10.1177/1745691623118110.

[47] Betthäuser BA, Bach-Mortensen AM, Engzell P. A systematic review and meta-analysis of the evidence on learning during the COVID-19 pandemic. Nature Human Behaviour. 2023;7(3):375–385. doi:10.17605/osf.io/u8gaz.

[48] Goldstein E, Lipsitch M, Cevik M. On the effect of age on the transmission of SARS-CoV-2 in households, schools, and the community. The Journal of Infectious Diseases. 2021;223(3):362–369. doi:10.1093/infdis/jiaa691.

[49] Viner RM, Mytton OT, Bonell C, Melendez-Torres G, Ward J, Hudson L, et al. Susceptibility to SARS-CoV-2 infection among children and adolescents compared with adults: a systematic review and meta-analysis. JAMA Pediatrics. 2021;175(2):143–156. doi:10.1001/jamapediatrics.2020.4573.

[50] Franco N, Coletti P, Willem L, Angeli L, Lajot A, Abrams S, et al. Inferring age-specific differences in susceptibility to and infectiousness upon SARS-CoV-2 infection based on Belgian social contact data. PLoS Computational Biology. 2022;18(3):e1009965. doi:10.1371/journal.pcbi.1009965.

[51] Sah P, Fitzpatrick MC, Zimmer CF, Abdollahi E, Juden-Kelly L, Moghadas SM, et al. Asymptomatic SARS-CoV-2 infection: A systematic review and meta-analysis. Proceedings of the National Academy of Sciences. 2021;118(34):e2109229118. doi:10.1073/pnas.210922911.

[52] Flook M, Jackson C, Vasileiou E, Simpson CR, Muckian M, Agrawal U, et al. Informing the public health response to COVID-19: a systematic review of risk factors for disease, severity, and mortality. BMC Infectious Diseases. 2021;21:1–23. doi:10.1186/s12879-021-05992-1.

[53] O’Driscoll M, Ribeiro Dos Santos G, Wang L, Cummings DA, Azman AS, Paireau J, et al. Age-specific mortality and immunity patterns of SARS-CoV-2. Nature. 2021;590(7844):140–145. doi:10.1038/s41586-020-2918-0.

[54] Angeli L, Caetano CP, Franco N, Coletti P, Faes C, Molenberghs G, et al. Assessing the role of children in the COVID-19 pandemic in Belgium using perturbation analysis. Nature Communications. 2025;16(1):2230. doi:10.5281/zenodo.14777392.

[55] Elbaz IM, El-Metwally H, Sohaly M. Viral kinetics, stability and sensitivity analysis of the within-host COVID-19 model. Scientific Reports. 2023;13(1):11675. doi:10.1038/s41598-023-38705-6.

[56] Gao S, Binod P, Chukwu CW, Kwofie T, Safdar S, Newman L, et al. A mathematical model to assess the impact of testing and isolation compliance on the transmission of COVID-19. Infectious Disease Modelling. 2023;8(2):427–444. doi:10.1016/j.idm.2023.04.005.

[57] Gebremeskel AA, Berhe HW, Atsbaha HA. Mathematical modelling and analysis of COVID-19 epidemic and predicting its future situation in Ethiopia. Results in Physics. 2021;22:103853. doi:10.1016/j.rinp.2021.103853.

[58] Angeli L, Caetano CP, Franco N, Abrams S, Coletti P, Van Nieuwenhuyse I, et al. Who acquires infection from whom? A sensitivity analysis of transmission dynamics during the early phase of the COVID-19 pandemic in Belgium. Journal of Theoretical Biology. 2024;581:111721. doi:10.1016/j.jtbi.2024.111721.

[59] Ratre YK, Vishvakarma NK, Bhaskar L, Verma HK. Dynamic propagation and impact of pandemic influenza A (2009 H1N1) in children: a detailed review. Current Microbiology. 2020;77(12):3809–3820. doi:10.1007/s00284-020-02213-x.

[60] Worby CJ, Chaves SS, Wallinga J, Lipsitch M, Finelli L, Goldstein E. On the relative role of different age groups in influenza epidemics. Epidemics. 2015;13:10–16. doi:10.1016/j.epidem.2015.04.003.

[61] Sekine T, Perez-Potti A, Rivera-Ballesteros O, Strålin K, Gorin JB, Olsson A, et al. Robust T cell immunity in convalescent individuals with asymptomatic or mild COVID-19. Cell. 2020;183(1):158–168. doi:10.1016/j.cell.2020.08.017.

[62] Burgess S, Ponsford MJ, Gill D. Are we underestimating seroprevalence of SARS-CoV-2? BMJ. 2020;370:m3364. doi:10.1136/bmj.m3364.

[63] Klinkenberg D, Backer J, Reusken C, Wallinga J. Seasonal variation in SARS-CoV-2 transmission in the Netherlands, 2020-2022: statistical evidence for a negative association with temperature. medRxiv. 2024; p. 2024–11. doi:10.1101/2024.11.28.24318154.

[64] Endo A, Group CCW, Uchida M, Liu Y, Atkins KE, Kucharski AJ, et al. Simulating respiratory disease transmission within and between classrooms to assess pandemic management strategies at schools. Proceedings of the National Academy of Sciences. 2022;119(37):e2203019119. doi:10.1073/pnas.220301911.

[65] Lasser J, Sorger J, Richter L, Thurner S, Schmid D, Klimek P. Assessing the impact of SARS-CoV-2 prevention measures in Austrian schools using agent-based simulations and cluster tracing data. Nature Communications. 2022;13(1):554. doi:10.5281/zenodo.4706876.

[66] Leng T, Hill EM, Holmes A, Southall E, Thompson RN, Tildesley MJ, et al. Quantifying pupil-to-pupil SARS-CoV-2 transmission and the impact of lateral flow testing in English secondary schools. Nature Communications. 2022;13(1):1106. doi:10.5281/zenodo.5898631.

[67] Munday JD, Sherratt K, Meakin S, Endo A, Pearson CA, Hellewell J, et al. Implications of the school-household network structure on SARS-CoV-2 transmission under school reopening strategies in England. Nature Communications. 2021;12(1):1942. doi:10.1038/s41467-021-22213-0.

[68] Boldea O, Alipoor A, Pei S, Shaman J, Rozhnova G. Age-specific transmission dynamics of SARS-CoV-2 during the first 2 years of the pandemic. PNAS nexus. 2024;3(2):pgae024. doi:10.1093/pnasnexus/pgae024.

[69] Rodiah I, Vanella P, Kuhlmann A, Jaeger VK, Harries M, Krause G, et al. Age-specific contribution of contacts to transmission of SARS-CoV-2 in Germany. European Journal of Epidemiology. 2023;38(1):39–58. doi:10.1007/s10654-022-00938-6.

## References

[1] PORDATA. Base de Dados Portugal Contemporâneo; 2024. Available from: https://www.pordata.pt/pt.

[2] Mistry D, Litvinova M, Pastore y Piontti A, Chinazzi M, Fumanelli L, Gomes MF, et al. Inferring high-resolution human mixing patterns for disease modeling. Nature Communications. 2021;12(1):323. doi:10.1038/s41467-020-20544-y.

[3] Backer JA, Mollema L, Vos ER, Klinkenberg D, Van Der Klis FR, De Melker HE, et al. Impact of physical distancing measures against COVID-19 on contacts and mixing patterns: repeated cross-sectional surveys, the Netherlands, 2016–17, April 2020 and June 2020. Eurosurveillance. 2021;26(8):2000994. doi:10.2807/1560-7917.ES.2021.26.8.2000994.

[4] Gabinetes da Secretária de Estado Adjunta e da Educação e do Secretário de Estado da Educação. Díario da República, 2.ª série — N.° 115 — 18 de junho de 2019; 2019. Available from: https://files.diariodarepublica.pt/2s/2019/06/115000001/0000200005.pdf.

[5] Viana J, van Dorp CH, Nunes A, Gomes MC, van Boven M, Kretzschmar ME, et al. Controlling the pandemic during the SARS-CoV-2 vaccination rollout. Nature Communications. 2021;12(1):3674. doi:10.1038/s41467-021-23938-8.

[6] Kislaya I, Gonçalves P, Barreto M, Sousa Rd, Garcia AC, Matos R, et al. Seroprevalence of SARS-CoV-2 infection in Portugal in May-July 2020: results of the first national serological survey (ISNCOVID-19). Acta Medica Portuguesa. 2021;34(2):87–94. doi:10.20344/amp.15122.

